# Supremacy of attention based convolution neural network in classification of oral cancer using histopathological images

**DOI:** 10.1101/2022.11.13.22282265

**Authors:** Bhaswati Singha Deo, Mayukha Pal, Prasanta K. Panigrahi, Asima Pradhan

## Abstract

**Introduction:** Oral cancer has grown to be one of the most prevalent malignant tumours and one of the deadliest diseases in emerging and low-to-middle income nations. The mortality rate can be significantly reduced if oral cancer is detected early and treated effectively.

**Objectives:** This study proposes an effective histopathological image classification model for oral cancer diagnosis using Vision Transformer deep learning based on multi-head attention mechanism.

**Methods:** The oral histopathological image dataset used in the study consists of 4946 images, which were categorized into 2435 images of healthy oral mucosa and 2511 images of oral squamous cell carcinoma (OSCC). In our proposed approach, along with Vision Transformer model eight pre-trained deep learning models known as Xception, Resnet50, InceptionV3, InceptionResnetV2, Densenet121, Densenet169, Densenet201 and EfficientNetB7 have been used for the comparative analysis. 90% of the images are used for training the models while the rest 10% of the images are used for testing purposes.

**Results:** Vision Transformer model achieved the highest classification accuracy of 97.78% in comparison to other considered deep learning models. Specificity, sensitivity and ROC AUC score are recorded as 96.88%, 98.74% and 97.74% respectively.

**Conclusion:** We found that our proposed Vision Transformer model outperforms compared to other pre-trained deep learning models, demonstrating a stronger transfer ability of the learning in histopathological image classification from the analysis of the obtained results. This method considerably lowers the cost of diagnostic testing while increasing the diagnostic effectiveness, and accuracy for oral cancer detection in patients of diverse origin.

## 1. Introduction

Oral cancer is indeed a fatal condition with a complex aetiology and a high death rate. With more than 377,700 cases recorded globally in 2020, the World Cancer Fund Research International claims that malignancies of the lip and oral cavity are the most common. According to (1), malignancies of the oral cavity and lip are the 11th and 18th most frequently occurring in both men and women. A well-formulated strategy is required for addressing oral cancer which includes early detection, risk factor management, and health literacy. Risk factors include consuming alcohol, smoking, having poor dental hygiene, being exposed to the human papillomavirus (HPV), geographical location, lifestyle, and ethnicity (2).

Squamous cell carcinoma (SCC) can develop from precancerous lesions such as erythroleukoplakia, oral leukoplakia, and verrucous hyperplasia (3). 90% of all oral cancers are SCCs (4). The most accurate way to diagnose oral cancer is through biopsy; however, this method is painful, and in cases of extensive or many lesions, selecting the appropriate site and size for surgical treatment of the biopsy sample can be challenging (5). Additionally, due to lesion variability, the prepared histology specimen may not accurately reflect the identification of the entire lesion. To achieve a successful cure, higher chances of survival, reduced death and morbidity rates, oral squamous cell carinoma (OSCC) must be detected early (6). The OSCC has a poor outlook with only a 50% average cure rate (7; 8). The accepted approach for diagnosing OSCC is tissue sample histopathological examination based on microscopy (9; 10). The clinical value of this diagnostic pathology approach is constrained by the histopathologists’ interpretation, which is frequently laborious and prone to error (11). Therefore, it is crucial to offer efficient diagnostic techniques to support pathologists in the evaluation and diagnosis of OSCC.

Because of the rise in processing power in recent years, deep learning algorithms have become the state-of-the-art in field of computer vision and image processing (12; 13; 14) owing to their strength arising from the accessibility of vast volumes of data. As a result, numerous investigations have been conducted to aid pathologists, particularly those employing deep learning techniques, especially convolutional neural networks (CNNs) for classification, medical image segmentation and localization (15; 16; 17). The transformer based on attention mechanism architecture (18; 19) has recently outscored convolutional neural networks.

Therefore, transformer architecture has emerged as the leading tool in the field of natural language processing (NLP). Transformers are built utilising a self-attention-based learning mechanism that learns the correlation between output and input patterns without relying on repetition. This makes it possible for transformer implementations to be quickly and effectively parallelized. In response to the popularity of transformers in natural language processing (NLP), transformer architecture was redesigned, which they referred as Vision Transfomer (ViT) (18). In the reshaped version, the transformer accepts a series of fixed-size image patches as input and treats them similar to how words are used for image classification in NLP application. ViT performed poorer in comparison to convolution neural networks when it was trained from scratch (CNNs). However, pretraining a ViT on a large amount of data and then performing transfer learning on a smaller dataset helped in outperforming CNN architectures in several computer vision applications such as image classification (18), semantic segmentation (20), and object detection (21; 22). Despite the success of vision transformers with natural images, there hasn’t been much exploration into the subject for medical imaging. Analysis of medical images and diagnosis are still regularly performed using CNN-based models. CNNs have some drawbacks in comparison to transformers, for example, convolution operations being difficult to recognize global information (23) and CNNs being unable to identify long-range relationships between various images available in healthcare database (24).

Some researchers have proposed transformer-based architectures in the field of medical imaging and diagnostics as a solution to the CNN issues (25; 26). A multi-modal classification approach for medical imaging was proposed by (24). In order to study both global and low-level features for an advantageous classification and image fusion technique, the authors merged CNN with transformer. According to the methodologies described here, ViT outperforms CNN-based models, although they do not take into account the fine-tuning and pretraining elements of transformers. To maximise their capabilities, transformers require a large amount of training data (18). However, in health sector large datasets aren’t always available (27). ViT struggles with a lack of inductive bias on small datasets after training that leads to a lack of generalizability (18).

It has been observed in the past that CNN-based architectures perform better after pretraining is done on publicly available huge image datasets such as ImageNet (28) and subsequently transfer learning on smaller medical datasets (29; 30). On that account, in this study, we evaluated the image classification ability of pre-trained transformers on an oral histopathological image dataset. To help with medical diagnosis even when experienced pathologists are not available, we suggest an automated technique that can identify between normal and OSCC patients. The classification model was also compared with eight pre-trained deep learning models and recently published studies in order to establish a benchmark for our findings.

## 2. Related works

### 2.1. Deep learning models

A novel strategy for combining the bounding box annotations from various doctors was put out in the literature (31). Deep Neural Network (DNN) was additionally employed to build automated systems that tackle this demanding task by producing difficult patterns. Two DL-based CV techniques were taken into consideration for automatic detection and classification of oral lesions since a primary collected data in this case is used.

Jeyaraj et al. (32) created a DL method that takes patient hyperspectral images into account for advanced and computeraided oral cancer diagnosis. The performance of the proposed regression-based partitioned DL strategy was assessed against other methods in terms of classifier accuracy, sensitivity, and specificity in order to validate it.

Using typical primary medical images, Jubair et al.(15) developed a lightweight DCNN for image classification of oral lesions, separating benign from malignant or potentially malignant lesions. The training and testing of the proposed technique used a data set of 716 medical images.

Instead of using digitized microscopic images for OSCC grade assessment, Das et al. (33) selected small patches from 156 WSIs and digitalized them at a 40X magnification. They conducted a comparison between the transfer learning strategy using the Alexnet, VGGNet, and ResNet models with that of the models trained from scratch. With an accuracy of 97.51%, the results demonstrated that convolutional neural networks (CNNs) created from scratch outperformed transfer learning models.

Popular CNNs like UNet and Inceptionv4 have shown significant promise in identifying and localising regions of abnormality on digital slides for oral squamous cell carcinoma (34; 35). With the aid of DL methods, Nanditha et al. (36) developed an automated system to examine oral lesions. In this study, an ensemble DL method was developed that combines the benefits of Resnet-50 and VGG-16. The augmented dataset of oral images was used to train this approach.

Unfortunately, even for highly skilled and experienced specialists, diagnosing oral cancer is still extremely difficult because histopathological images provides diversified morphological features. Because of this, diagnosing oral cancer using conventional techniques is energy and time consuming, so it becomes difficult to use a standardised method to classify oral cancer images as normal or OSCC. CNN’s performance has seen significant improvements over time due to the efforts of numerous researchers. Contrarily, the CNN model only looks at the relationship between spatially located adjacent pixels in the interested area determined by the filter size. As a consequence, it is challenging to determine connections with far off pixels. Recently, efforts have been made to use attention processes to solve this problem, which is capable of identifying long range correlation.

### 2.2. Attention mechanism

The attention mechanism is the most recent state-of-the-art development in computer vision problems (37; 38; 39). The attention strategy rejects redundant features in favour of valuable features. It has taken the place of the conventional CNNs methods, which concentrate on all of the features of the input image, paying attention to both significant and repetitive information, and thus giving wrong predictions. Without increasing the model’s computing cost, attention mechanisms can enhance interpretation and maximise classifier performance.

In work of (40), SENet was used to retrieve relevant image channel weights, highlighting how removing data redundancy might improve a deep learning model’s ability to better classify images. With minimally increased computational complexity, SENet considerably reduced prediction error. The issue of SENet’s growing model complexity was brought up by ECA-Net (41). ECA-Net eliminates the dimensionality contraction process in the fully connected layer by using cross-channel connections with 1D convolutions. They can thereby increase speed while reducing model complexity. Zhang et al. (41) proposed a novel ‘Shuffle Attention’ module to address the computation complexity. They improved classification accuracy over ImageNet-1k by more than 1.34% by subjecting the input features to spatial and channel attention. Because it collects data on microscopic lesions, this technique performs significantly better in the medical field. In (42), a deep network for concurrent microvessel and nerve segmentation in frequently used hematoxylin and eosin (H&E) stained histopathological images is investigated. The network has an encoder-decoder architecture embedded with feature attention blocks.

### 2.3. Vision Transformers

Transformers (43; 19) are deep neural network models based on attention mechanisms that were initially developed for natural language processing (NLP) applications. After reaching breakthrough performance in natural language processing, the vision community studied its applicability in computer vision challenges to make use of its capacity to recognize far-off dependencies within an image. (44). In comparison to state-of-the-art CNN in image classification evaluations, the Vision Transformer (ViT) performs well. It is one of the favourable attempts to exploit Transformer specifically on images (18; 45; 46). Despite having better performance, it has an easy-to-use modular framework that allows for wide-ranging application in multiple tasks with minimal modification. The image recognition transformer proposed by Chen et al. (47) was one of the prospective multi-task architectures for resolving a variety of computer vision issues.

Transformers are used for computer vision as a result of their enormous success in the field of natural language processing. One of the most widely used image classification transformers is OpenAi Image GPT abbreviated as iGPT (48), that uses ImageNet for model training and GPT to generate images. However, iGPT suffers from low image quality and high computational requirements. Google ViT (18) is an image categorization tool that turns images into patches and feeds them to the original transformer architecture. Facebook Data Efficient Transformer (DeiT) (49) is having a similar framework as Vision Transformer and knowledge distillation is used to enhance model training. CNN serves as the instructor model in this system. A transformer-based approach for segmenting medical images is called Segtran (50). By exploiting the endless transformer’s receptive fields, it contextualizes features. Segtran produces good segmentation results because it can perceive both the overall view and the fine details.

### 2.4. Research gaps and motivation

1. As was previously stated, a great number of people are affected by oral cancer, particularly in rural and underdeveloped countries, which are characterised by a variety of variables including lifestyle, ethnicity, alcohol usage, and a lack of adequate medical care. To be treated, oral cancer needs to be discovered as soon as feasible.
2. The assessment of histopathological images is the most used technique for diagnosis. However, it depends on the pathologist’s ability to interpret. A quick, accurate model with generalizing capability is required to identify the problem.
3. According to recent research, conventional neural networks (CNN) perform notably better than conventional methods since they automatically extract features from the input image. In contrast, the spatial connection between neighbouring pixels in the focused area determined by the filter size is the focus of the CNN model. It is therefore difficult to establish relationships with distant pixels. To address this issue, attention mechanisms have recently been tried.
4. There are very few publicly accessible imaging datasets for oral cancer. It is well known that deep learning neural networks require a significant quantity of data for excellent classification accuracy. The Vision Transformer approach has employed transfer learning to overcome this issue.
5. Few authors have also used attention mechanisms that have been shown to be more structured and accurate at extracting relevant features from images than traditional models for deep learning in medical image classification purpose. However, there has been minimal focus on more modern multi head attention strategies, which in computer vision tasks have been demonstrated to be superior to single-head attention processes.

### 2.5. Research contributions

Here is a list of this paper’s main contributions:

1. The proposed work emphasized on medically relevant, explainability-driven visuals to highlight the key areas critical to input image for binary classification of images based on Vision Transformer.
2. Transformer-based feature extraction framework is used as a backbone of the proposed binary classification framework.
3. We have used a publicly available oral cancer histopathological image dataset (51) in order to undertake a thorough and reliable binary image classification analysis.
4. We have compared the performance ability of our proposed model with eight pre-trained image classification deep learning models.
5. This study did not take into account the method of enhancing the quality of histopathological images as conducted in previous studies for visual improvements because our proposed approach divides the input images into patches, then flattens each patch into a separate feature vector.

The remaining part of this work is written out as follows: section “Proposed Method” discusses details of the Vision Transformer, section “Pre-trained deep learning models for comparison” gives a brief description about eight considered deep learning (DL) models for comparative purpose, section “Data and materials” includes description of dataset, preprocessing, training criteria, data augmentation techniques and implementation details, section “Results and Discussion” presents the results of the proposed method along with considered DL models used for comparison. Finally, section “Conclusion” summarizes the work and outlines the future scope of our proposed approach.

## 3. Proposed Method

This section discusses in detail the proposed classification methodology for oral cancer histopathological image classification as shown in Figure 1. We used the conventional Vision Transformer (ViT) model trained on the ImageNet dataset (28). This is done in order to assess the efficacy of ViT’s transfer learning from non-medical images to medical images. The ViT model we are utilising was greatly influenced by (18) and is closely connected to the original Transformer (19). Figure 2 depicts the ViT model’s architecture in detail, which is used to categorize oral cancer images. The input images are divided into two dimensional patches of fixed size, flattened, and integrated with position embeddings before being fed in succession to the ViT model. The transformer encoder is made up of repeating blocks with layers of normalization, multi-head attention, and multilayer perceptron (MLP) layers. The encoded feature vector is mapped to one of the output classes by the MLP classification head, which is coupled to the encoder blocks’ output. We contrast the effectiveness of ViT’s transfer learning with that of CNN-based architectures like Xception (52), ResNet-50 (12), InceptionV3 (53), InceptionResnetV2 (54), Densenet121 (55), Densenet169 (55), Densenet201 (55), and EfficientNetB7 (13). Additionally, transfer learning from ImageNet is used to train these CNNs (28). Accuracy, precision, recall, and F1-score are among the evaluation measures used to assess the performance of ViT model and CNNs.

**Figure 1:**
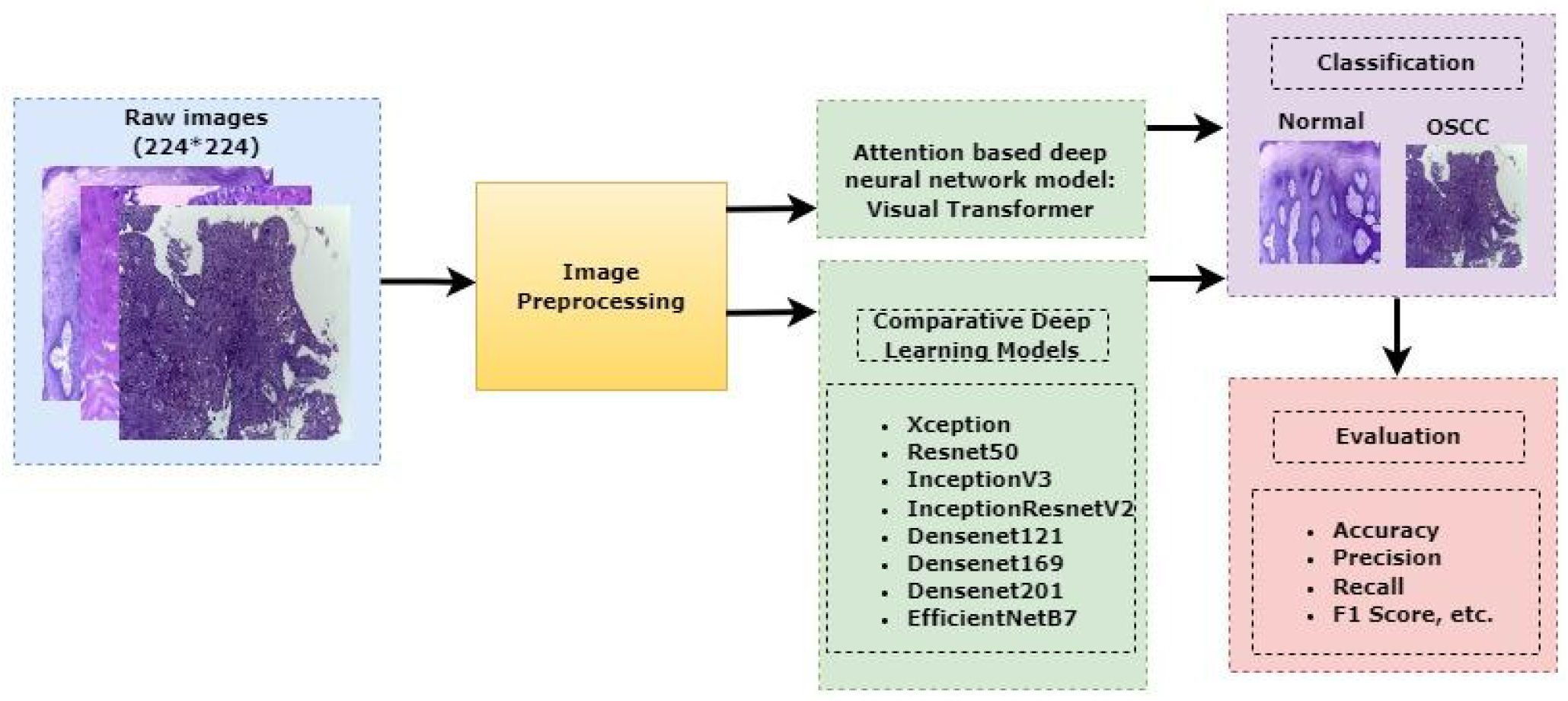
Proposed classification methodology for oral cancer image classification

**Figure 2:**
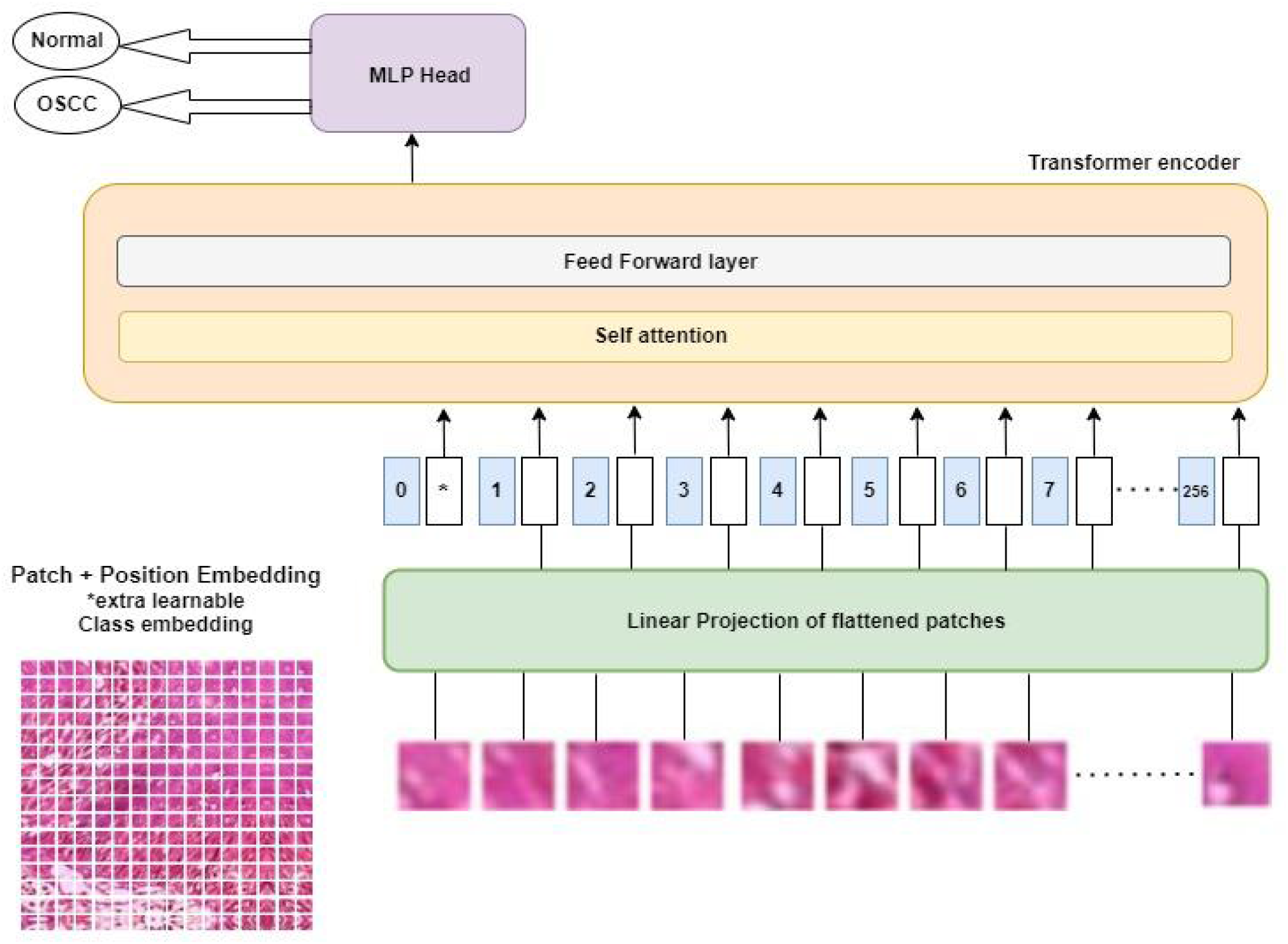
Details of Vision Transformer model for oral cancer detection

### 3.1. Description of Transformer Encoder

The Vision Transformer’s encoder block fetches histopathological images that are divided into 14 × 14 pixel patches. After positional encoding, the patches are encoded in the form of a feature matrix represented as X. Image’s spatial structure is preserved owing to positional encoding. The correlations between patches are described using a selfattention method. This method is based on three embeddings termed as query, key and value. Query, key and value are abbreviated as Q, K and V respectively. It is defined as follows:

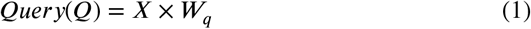

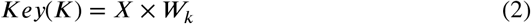

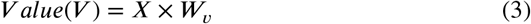

Patch features are projected onto Q, K, and V embeddings using *W*_*q*_, *W*_*k*_, and *W*_*v*_ respectively. Self attention technique and features based on attention weights comprises the majority of the ViT encoder pipeline’s operations. The dependencies between patches are modelled using the selfattention method. This is how the self-attention approach functions: Initially, the dot product between Q and K is used to determine the correlation between patches embeddings as shown below:

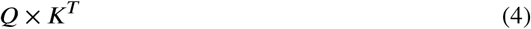

The scores are scaled down as shown below. Since multiplying values can have large magnitude, this scaling factor allows for gradients that are more steady:

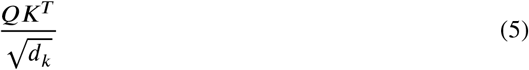

The softmax layer transforms the similarity measure between Q and K into a probability distribution. As a result, the model is more aware of which patch to concentrate on.

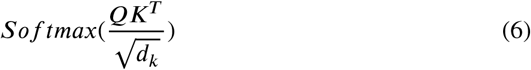

Attention weights computed for each of the features in the preceding step are known as self attention scores. This score decides the weight of the value embeddings.

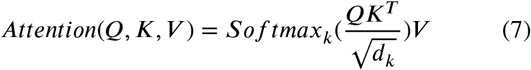

The calculation of attention scores as indicated in the preceding Eq. (7) is displayed in Figure 3. MatMul is short for matrix multiplication here. Concatenation is referred to by the acronym concat.

**Figure 3:**
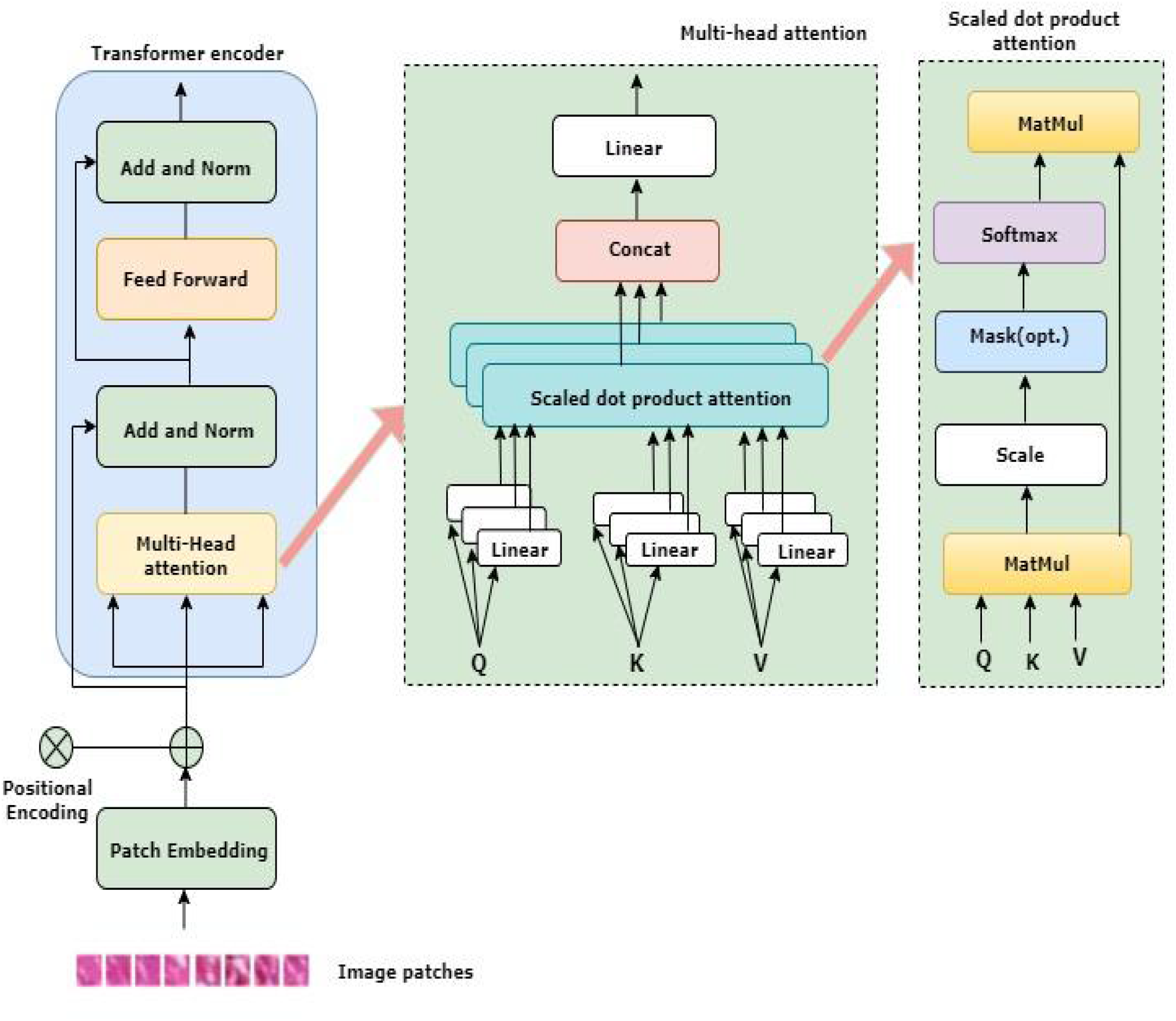
Transformer encoder for processing oral cancer histopathological image patches with multihead attention and scale dot product attention module.

**Figure 4:**
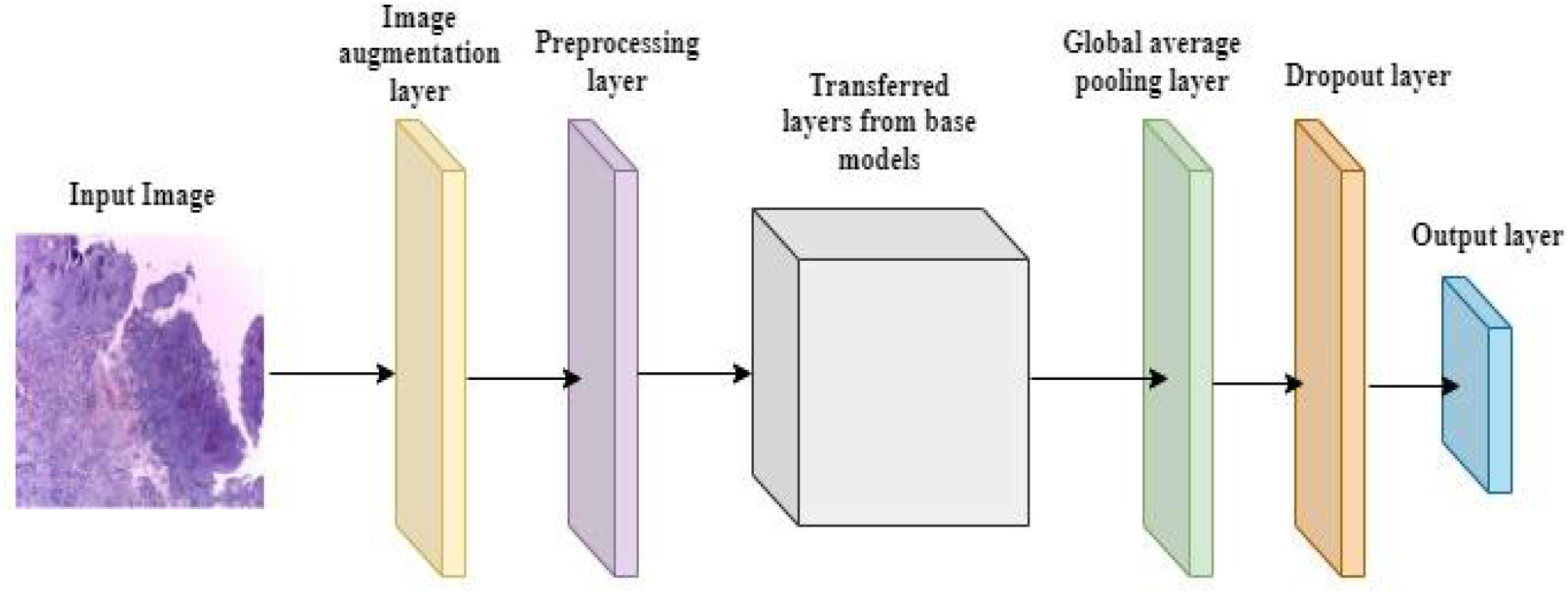
Convolutional neural network of the considered models for comparative analysis with Visual Transformer using oral cancer histopathology images

### 3.2. Description of Multi-headed Attention block

Each self attention operation serves as a head in multiheaded attention mechanism, and each head tries to learn something unique, improving the encoder model’s representation power. While computing multi-headed attention, Q, K and V are split into N number of vectors before self-attention is applied, as illustrated in Figure 3. The split vectors then go through the self-attention procedure one at a time. Each head represents a step in the development of self-awareness. So every head produces an output vector, which is then concatenated into a vector and passed through the final linear layer.

## 4. Pre-trained deep learning models for comparison

In this study, we compared the effectiveness of the proposed Visual Transformer model using eight pre-trained deep learning (DL) models for the classification of oral cancer histopathological images. Here are some details on the pre-trained DL models that were chosen: Xception (52), Resnet50 (12), InceptionV3 (53), InceptionResnetV2 (54), Densenet121 (55), Densenet169, (55), Densenet201 (55), EfficientNetB7 (13).

### 4.1. Xception

Xception, which refers to “extreme inception” pushes the fundamental ideas of Inception to the utmost (52). In Inception, the initial input was compressed using 1×1 convolutions, and from each one of those input image, we used various types of filters on each depth space. Just the opposite occurs with Xception. Instead, it applies the filters to each depth map individually before using 1×1 convolution to compress the input space all at once. The feature extraction backbone of the network in the Xception architecture is composed of 36 convolutional layers. The Xception architecture can be summed up as a linear stack of residually connected depthwise separable convolution layers. This makes defining and changing the architecture relatively simple.

### 4.2. Resnet50

Classification, auto-encoding, and images are integrated by a traditional deep convolutional neural network known as ResNet (12). The 2015 ImageNet Large Scale Visual Recognition Challenge (ILSVRC) was won by this method, which used feature transmission to reduce the vanishing gradient problem and allowed the training of far deeper networks than those previously used. It is widely known that adding layers on top of each other ineffectively increases network depth. Because of the vanishing gradient problem, when gradients are continuously multiplied as they are backpropagated to earlier layers, the gradient in deep networks may become very small. As a result, as the network goes deeper, its performance becomes saturable or even starts to degrade quickly.

ResNet introduced the skip connection initially. Figure 5 shows the skip connection. The skip connection connects activations from one layer to the next by skipping over some intermediary levels. As a result, a block is leftover. These extra blocks are heaped and used to construct resnets. The strategy adopted by this network is to let the network fit the residual mapping rather than having layers learn the underlying mapping. The benefit of this kind of skip link is that it enables regularisation to avoid any layers that reduce architecture performance. This means that when training an extremely deep neural network, disappearing or growing gradients are not a concern.

**Figure 5:**
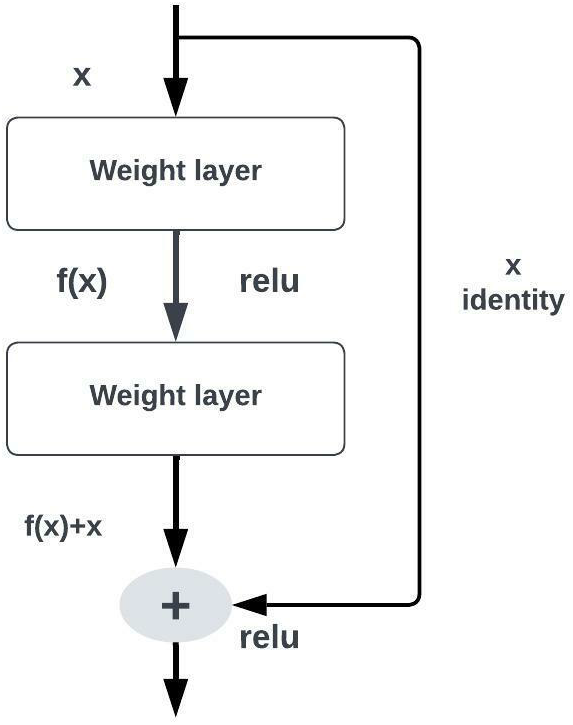
Exemplary structure of skip connection used in Resnet50 model

### 4.3. InceptionResnetV2

A convolutional neural network known as Inception-ResNet-v2 expands on the Inception group of architectures while incorporating residual connections. It replaces the filter concatenation step of the Inception model (54).

### 4.4. InceptionV3

On the ImageNet dataset, it has been demonstrated that the image recognition model Inception v3 can achieve higher than 77.9% accuracy. The model is the result of numerous concepts that have been established by various researchers over time. ‘Rethinking the Inception Architecture for Computer Vision’ by Szegedy et al. (53) served as its foundation. Concatenations, convolutions, max pooling, dropouts, average pooling, and fully connected layers are some of the symmetric and asymmetric building components that make up the model itself. The model makes considerable use of batch normalisation, which is also applied to the activation inputs. To calculate loss, Softmax is used.

### 4.5. Densenet121/169/201

The feature maps of the current layer and all previous layers are combined to generate each layer in (DenseNets 55). As a result, these networks are effective in terms of computation and memory utilisation, have rich extracted features of the input images, and are small (i.e., have fewer channels). Shorter connections between the layers nearest to the input and the layers nearest to the output enable faster, deeper, and more accurate training of CNN.

A feed-forward connection is made between each layer of the dense convolutional network (DenseNet). Each layer uses the feature-maps of all the layers before it, and all succeeding layers utilise the feature-map of the layer in use as their input. DenseNet, which further improves feature propagation, increases feature reuse, and significantly lowers the number of parameters, prevents the vanishing gradient problem and mitigates its impacts.

### 4.6. EfficientNetB7

EfficientNetB7 is a non-repetitive, non-linear neural network search that optimizes floating point operations per second (FLOPS) and accuracy by balancing resolution, network depth and breadth. Seven flipped residual blocks, each with its own parameters, are used in the architecture. These blocks employ swish activation, squeeze, and excitation blocks (13).

## 5. Data and Materials

### 5.1. Dataset Description

The image set (51), which is accessible to the public, is used to retrieve microscopic images stained with H&E. There are two types of subjects in the considered oral cancer dataset: the patients having oral cancer and the healthy subjects. The suggested approach assesses and predicts oral cancer using this dataset. While table 2 shows the entire dataset classes, figure 6,7 shows samples from both dataset categories.

**Table 1.** A brief introduction to the deep learning models used along with the Vision Transformer for a comparative study in medical image classification

| Model | Ref. | No. of parameters | Major remarks |
| --- | --- | --- | --- |
| Xception | (52) | 22.9M | (a)It is InceptionV3's improved version.<br>(b)It makes use of separable convolutions in depth. |
| Resnet50 | (12) | 25.6M | (a)There is a new block called residue.<br>(b)The issue of accuracy rapidly declining as a result of an increase in network layers has been resolved. |
| InceptionV3 | (53) | 23.9M | (a)It makes use of a better Inception Module.<br>(b)Small convolutions are used to introduce the concept of factorization. |
| InceptionResnetV2 | (54) | 55.9M | (a)The ResNet connection and the Inception block are combined. |
| Densenet121 | (55) | 8.1M | (a)When two layers have the same feature map size, a direct connection between them is established.<br>(b)Hundreds of layers are possible at this scale. |
| Densenet169 | (55) | 14.3M |  |
| Densenet201 | (55) | 20.2M |  |
| EfficientNetB7 | (13) | 66.7M | (a)Reduces parameter size and increases accuracy. |

**Table 2.** OSCC biopsy data instances

| Classes | Images |
| --- | --- |
| Normal | 2435 |
| OSCC | 2511 |

**Figure 6:**
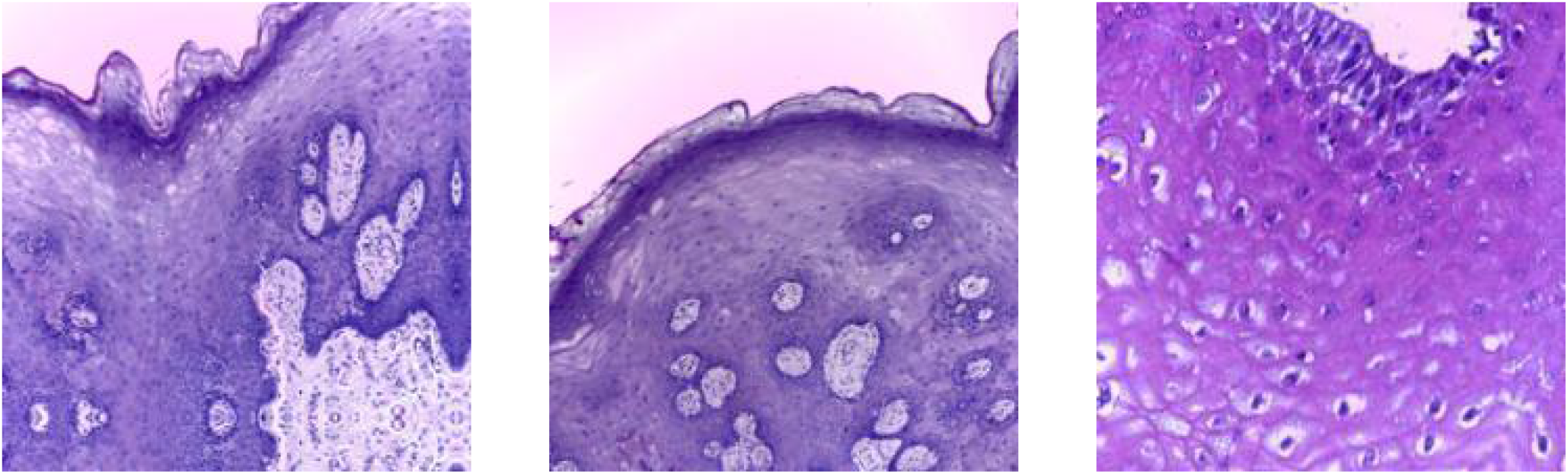
Sample Images from the Normal subjects

**Figure 7:**
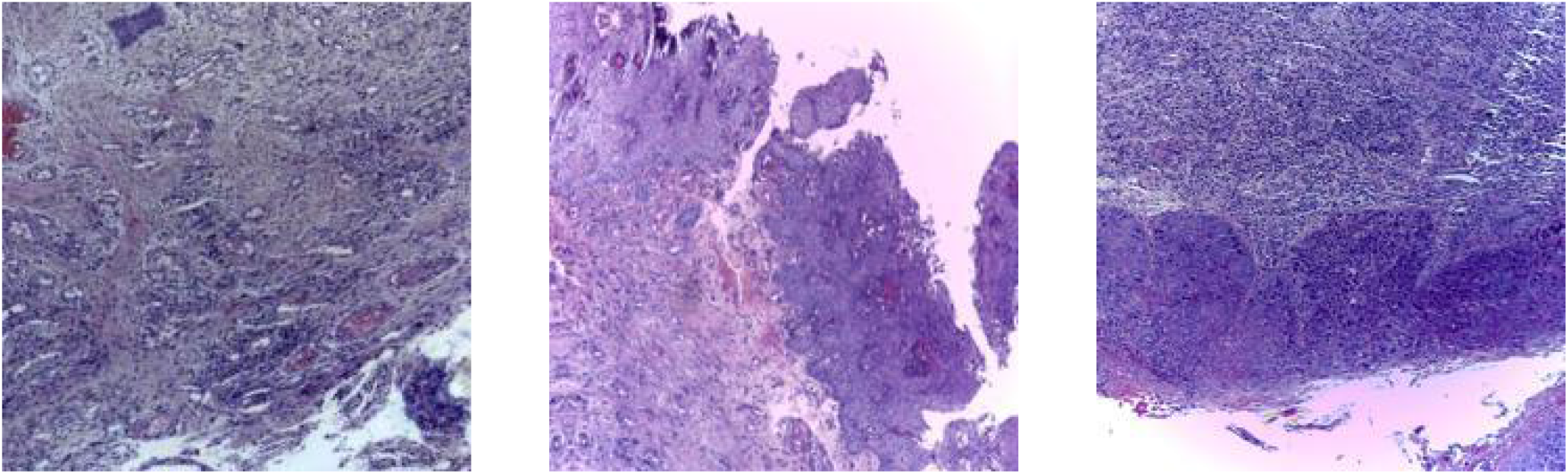
Sample Images from the OSCC patients

### 5.2. Preprocessing

Images in the dataset have a 224 × 224 pixel resolution. The input image’s pixel values are scaled between 0 and 1, and each input channel is normalised in reference to the ImageNet dataset.

### 5.3. Training Criteria

90% of the images were chosen for training, while 10% were chosen for testing. In vision transformer model, 4451 images were used for training purpose and 495 images were used to test the performance of the model. While in pretrained DL models, 4450 images were used to train the model, and 496 images were used in test criteria. This information on the training and testing of the models is shown in table 3.

**Table 3.** Criteria for training, validation and test images

|  | Images used |  |
| --- | --- | --- |
|  | DL models | Vision Transformer |
| Training | 4450 | 4451 |
| Test | 496 | 495 |

### 5.4. Data Augmentation

Increasing the number of images in the dataset results in a larger dataset, which is what is meant by “Image augmentation”. A vast number of images that are similar to one another could be produced by using Image augmentation. The dataset grows in size as a result. The model is forced to generalise when more data is introduced because it is unable to overfit all the samples. In this way, image augmentation minimises and even eliminates overfitting. We utilise the ImageDataGenerator function provided by the Keras deep learning toolbox to generate sets of tensor image data with relevant data augmentation. We want to ensure that each time our network is trained, it sees new iterations of our data using this type of data augmentation. The ImageDataGenerator receives a batch of input images before randomly rotating, flipping, standardizing, changing the brightness of, shifting, and performing other operations on every image in the batch. By setting “Random Flip = Horizontal,” the images are rotated horizontally. Image rotation is a popular technique for augmentation, and the ImageDataGenerator class rotates images to a certain extent at random. Additionally, we select “Random height = 0.2”, which establishes the maximum percentage of total height by which the image may be arbitrarily shifted upward or downward. By setting “Random zoom = 0.2 to 0.2,” images are zoomed in by a height factor of 0.2 and width factor of 0.2. The batch that underwent random alteration is then returned to the caller function. Each of these variables, together with their values, are shown in table 4.

**Table 4.**
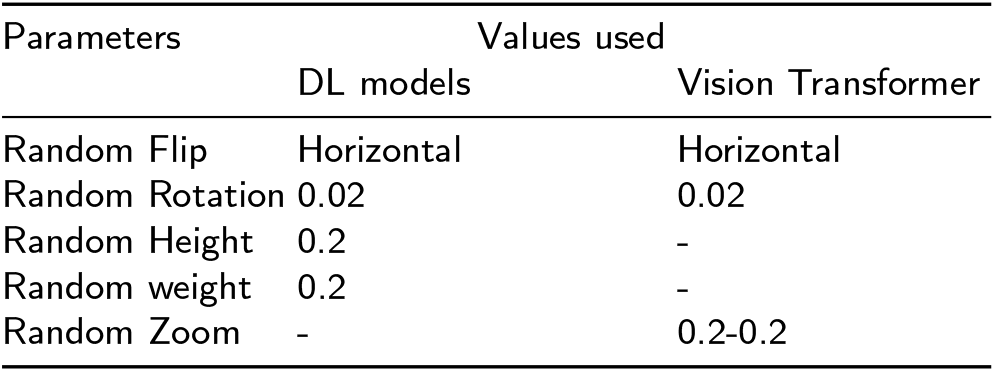
Parameters of Image Augmentation

### 5.5. Implementation

All studies related to this article were carried out using Python 3.7.6, TensorFlow 2.7.0, and Keras 2.7.0 on a standard PC with Intel(R) Iris(R) Xe graphics. A 2.40 GHz Intel(R) Core(TM) i5-1135G7 processor and 16.0 GB of RAM are also features of the laptop.

## 6. Results and Discussion

We first detailed the various evaluation metrics being used in our proposed approach, and finally we discussed how to adjust the hyperparameters and analyze the performance of Vision Transformer along with the considered DL models on the oral cancer histopathological image dataset.

### 6.1. Evaluation Metrics

The contents of the confusion matrix, also known as the error matrix or contingency table, are crucial to the overall performance of our proposed approach. Four terms—True Positive (TP), False Positive (FP), False Negative (FN), and True Negative—are included in this evaluation matrix (TN). In our illustration, the TP stands for images that were correctly identified as OSCC, whereas the FP stands for images that were incorrectly identified as normal. While the FN stands for OSCC class images that were incorrectly labelled as normal and the TN for correctly classified normal images. Using the Python scikit-learn module, seven performance measures based on the confusion matrix: precision, sensitivity, classification accuracy, F1-score, cohen kappa score, matthews correlation coefficient and brier score loss were used to assess the classification performance of our proposed model on the test images. These performance metrics can be computed using the formulas below:

1. Precision: The precision is calculated as the ratio of positive samples that were correctly classified to all samples that were classified as positive (either correctly or incorrectly). The precision gauges how well the model classify a sample as positive.

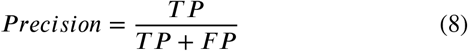
2. Sensitivity: Sensitivity calculates a model’s degree of completeness. It shows the proportion of images correctly identified as OSCC compared to all OSCC images.

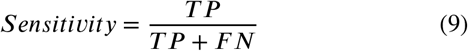
3. Accuracy: It measures the prediction performance and is expressed as the proportion of correctly identified images to all test images.

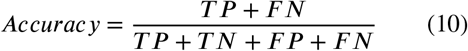
4. F1-Score: The harmonic mean of the model’s precision and recall is known as the F1 score. It is a method of optimizing the model towards either recall or precision.

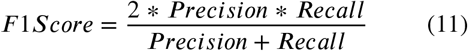
5. Cohen kappa score: It is calculated by the formula stated below as:

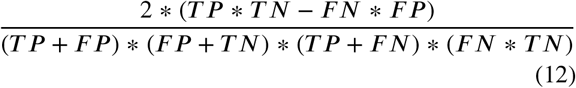
6. Matthews correlation coefficient: It is a metric used to evaluate the classification model’s effectiveness.

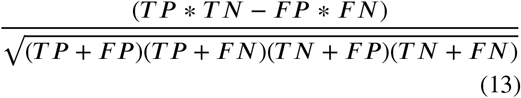
7. Brier score loss: The mean squared difference between the actual outcome and predicted probability is measured by the brier score.

### 6.2. Results and discussion of the proposed Vision Transformer model

In this section, we will discuss the results of the ViT model along with the hyperparameters being used for the proposed model.

#### 6.2.1. Hyperparameter tuning of Vision Transformer

We selected sparse categorical crossentropy as a loss function for our binary classification task. AdamW optimizer with weight decay of 0.0001 and learning rate of 0.001 is used for hyper-parameter optimization over the course of 100 epochs. We have used a patch size of 14×14 with a drop out of 0.5 and 4 heads in the encoder architecture. Batch size of 32 is used for best performance.

#### 6.2.2. Discussion of Vision Transformer model results

For the analytical results of Vision Transformer model on the histopathological oral cancer dataset, we have utilized a model pre-trained on the ImageNet dataset. We have used Vision Transformer as the base model and an additional softmax classifier on top of it to categorize images into normal and OSCC classes. The training and validation accuracy and loss curves are plotted over 100 epochs as displayed in figure 8a and 8b. The confusion matrix (CM) for Vision Transformer is also calculated to help in understanding the results. It displays the number of true positives (TP), false positives (FP), true negatives (TN), and false negatives (FN) as shown in figure 8c. It is inferred from the CM that FP and FN are very less in number as compared to TN and TP, thus showing correct predictions of images into normal and OSCC classes. After training and validation, we obtained the ROC (receiver operating characteristic) curve for the model, which shows performance exceeding 0.5 and an AUC (area under the curve) value of 0.98 as shown in Figure 9a. The precision-recall curve in figure 9b shows an average precision of 0.96.

**Figure 8:**
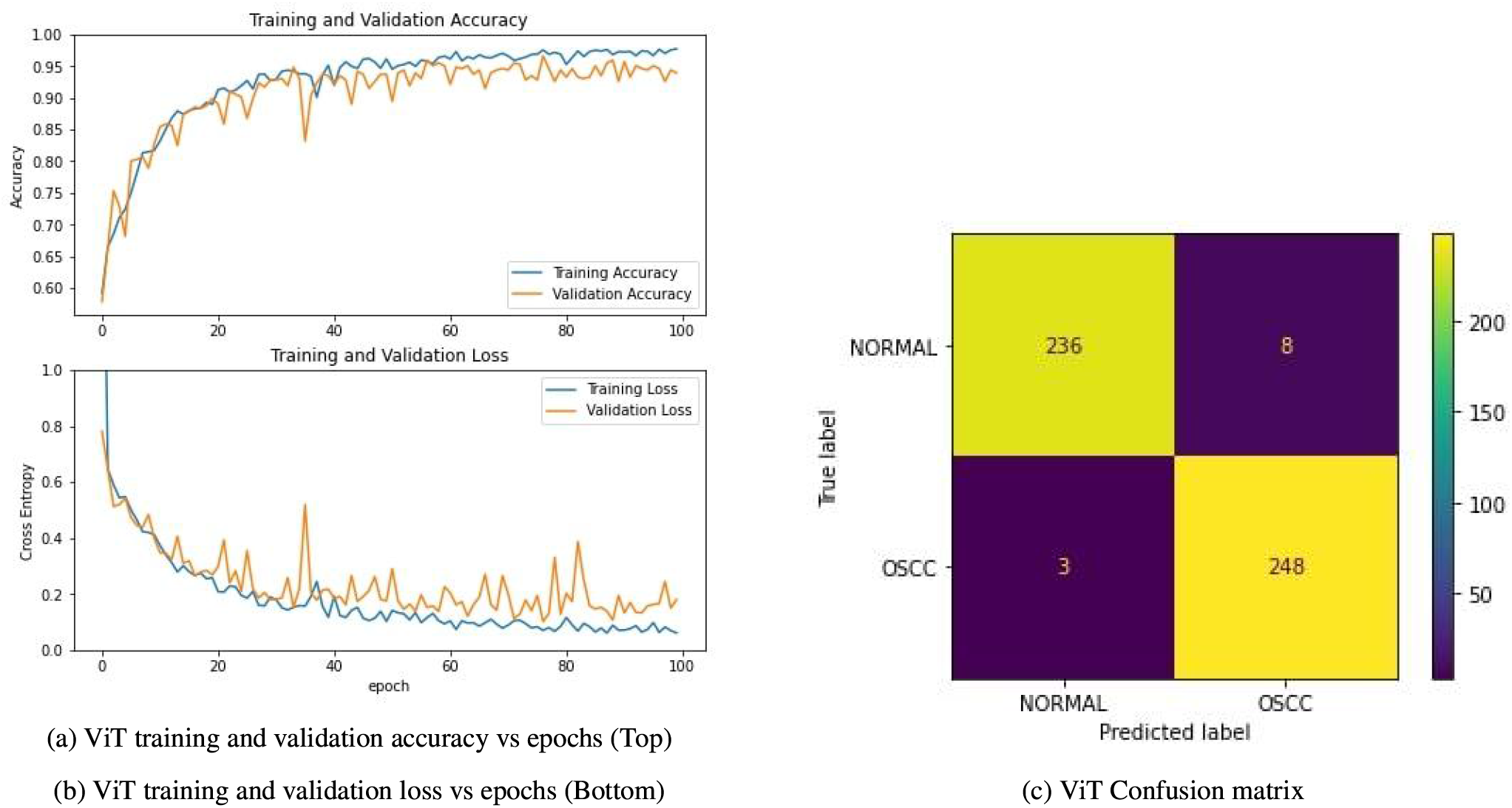
(a)Accuracy plot of ViT model training over 100 epochs(b)Loss plot of ViT model training over 100 epochs(c)Confusion matrix shows that there are 236 true negatives, 8 false positives, 3 false negatives and 248 true positives

**Figure 9:**
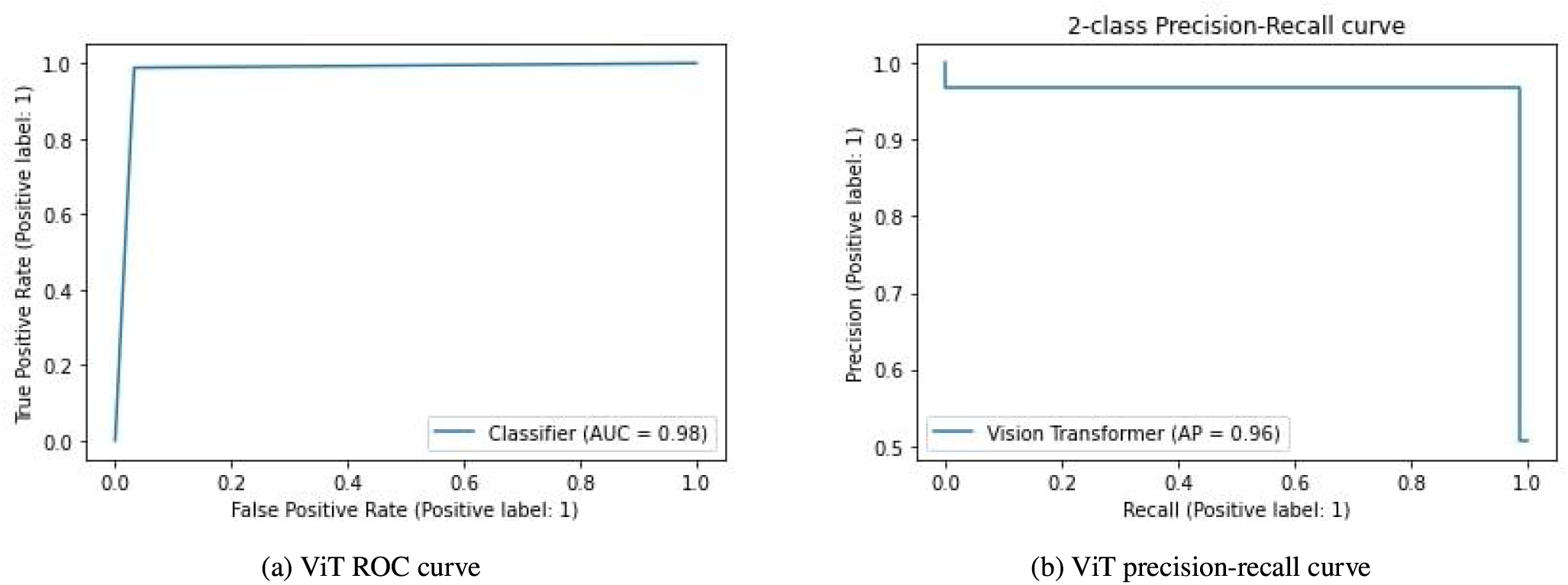
(a)ROC curve for the proposed ViT model. The area under the curve is calculated as 0.98(b)The precision-recall curve is constructed by plotting the precision against the recall for the ViT model. The average precision value is 0.96 which shows that our proposed model gets more correct predictions.

The overall model performance metrics are shown in table 6, 7, and 8. Table 9 shows a comparative analysis of our proposed work with previous research work.

**Table 5.** Hyperparameters used in the considered deep learning models and proposed Vision Transformer

| Hyperparameters | Considered DL models for comparison | Proposed Vision Transformer |
| --- | --- | --- |
| Optimizer | Adam | AdamW |
| Learning Rate | 0.001 | 0.001 |
| Weight decay | - | 0.0001 |
| Loss Function | Binary Cross-entropy | SparseCategoricalCrossentropy |
| Batch Size | 32 | 32 |
| Epochs | 100 | 100 |
| Dropout | 0.2 | 0.5 |
| Input size | $224 \times 224 \times 3$ | $224 \times 224 \times 3$ |
| Patch size | - | $14 \times 14 \times 3$ |
| Patches per image | - | 256 |

**Table 6.** Oral cancer image classification summary report

| Models | Class | Precision | Recall | F1 score |
| --- | --- | --- | --- | --- |
| Xception | Normal | 0.89 | 0.89 | 0.89 |
|  | OSCC | 0.89 | 0.89 | 0.89 |
| Resnet50 | Normal | 0.93 | 0.86 | 0.89 |
|  | OSCC | 0.88 | 0.93 | 0.90 |
| InceptionV3 | Normal | 0.85 | 0.82 | 0.84 |
|  | OSCC | 0.83 | 0.86 | 0.85 |
| InceptionResnetV2 | Normal | 0.90 | 0.82 | 0.86 |
|  | OSCC | 0.84 | 0.91 | 0.87 |
| Densenet121 | Normal | 0.92 | 0.69 | 0.79 |
|  | OSCC | 0.76 | 0.94 | 0.84 |
| Densenet169 | Normal | 0.91 | 0.77 | 0.83 |
|  | OSCC | 0.80 | 0.92 | 0.86 |
| Densenet201 | Normal | 0.92 | 0.84 | 0.88 |
|  | OSCC | 0.86 | 0.93 | 0.89 |
| EfficientNetB7 | Normal | 0.90 | 0.83 | 0.87 |
|  | OSCC | 0.85 | 0.91 | 0.88 |
| **Vision Transformer** | **Normal** | **0.99** | **0.97** | **0.98** |
|  | **OSCC** | **0.97** | **0.99** | **0.98** |

**Table 7.** Oral cancer image classification model performance accuracy metrics

| Models | Accuracy | Specificity | Sensitivity | ROC AUC score |
| --- | --- | --- | --- | --- |
| Xception | 88.70% | 88.87% | 88.52% | 89.00% |
| Resnet50 | 89.92% | 87.69% | 92.54% | 90.00% |
| InceptionV3 | 84.07% | 83.14% | 85.10% | 84.00% |
| InceptionResnetV2 | 86.69% | 83.94% | 90.09% | 87.00% |
| Densenet121 | 81.85% | 75.80% | 92.31% | 82.00% |
| Densenet169 | 84.68% | 80.34% | 90.78% | 85.00% |
| Densenet201 | 88.71% | 85.77% | 92.34% | 89.00% |
| EfficientNetB7 | 87.30% | 84.87% | 90.22% | 87.00% |
| **Vision Transformer** | **97.78%** | **96.88%** | **98.74%** | **97.76%** |

**Table 8.** Oral cancer image classification model performance evaluation metrics

| Models | Cohen kappa | Matthews correlation coefficient | Brier loss |
| --- | --- | --- | --- |
| Xception | 0.774 | 0.774 | 0.113 |
| Resnet50 | 0.798 | 0.799 | 0.101 |
| InceptionV3 | 0.681 | 0.681 | 0.159 |
| InceptionResnetV2 | 0.733 | 0.736 | 0.133 |
| Densenet121 | 0.636 | 0.657 | 0.181 |
| Densenet169 | 0.693 | 0.701 | 0.153 |
| Densenet201 | 0.774 | 0.777 | 0.101 |
| EfficientNetB7 | 0.746 | 0.748 | 0.127 |
| **Vision Transformer** | **0.956** | **0.956** | **0.022** |

**Table 9.** Comparative analysis with previous research

| Research work | Classification method | Dataset source | Dataset ref. | Classification accuracy |
| --- | --- | --- | --- | --- |
| (58) | Resnet101 | Private | Not available | 78.30% |
| (15) | EfficientNet-B0 | Private | Not available | 85.00% |
| (59) | Deep learning | Private | Not available | 88.30% |
| (60) | Alexnet | Public | (51) | 90.06% |
| **Current study** | **Proposed method** | **Public** | **(51)** | **97.78%** |

### 6.3. Results and discussion of the considered deep learning models

In this section, we will discuss the results of the deep learning models being considered for comparison with the proposed model along with its hyperparameters.

#### 6.3.1. Hyperparameter tuning of the considered DL models

Neural networks are powerfully capable of learning relationships between their inputs and outputs on their own, as demonstrated by (56). However, some of these connections might be the result of sampling noise, in which case they might be more prevalent during training but not in the real test dataset. This challenge might lead to overfitting problems that would make it more difficult for a deep learning model to predict (56). We closely followed the hyperparameter tweaking procedure in order to get the generalised performance of our suggested methodology.

The best hyperparameters were selected using the procedure described below: First, we selected binary cross-entropy as a loss function for our binary classification task. Then, over the course of 100 epochs, optimization was carried out using the Adam (adaptive moment estimation) approach (57) during the training process. The reduction of the generalization gap between training loss and validation loss was our main objective during model training, and we found that a batch size of 32 and a learning rate of 0.001 worked well together. Additionally, we used a dropout of 0.2 to prevent the model from becoming overfitted during training (36). Then, based on which model had the lower validation loss, we saved its weights. Notably, we adhered to the original Xception, Resnet50, InceptionV3, InceptionResnetV2, Densenet121/169/201, EfficientNetB7 architectures’ descriptions of convolutional filters, padding, pooling and strides. All of the ideal values for the hyperparameters utilised in this research are shown in table 5.

#### 6.3.2. Discussion of Xception model results

For the analytical results of Xception model on the histopathological oral cancer dataset, we have utilized a model pre-trained on the ImageNet dataset. We have used Xception as the base model and an additional softmax classifier on top of it to categorize images into normal and OSCC classes. The training and validation accuracy and loss curves are plotted over 100 epochs as displayed in figure 10a and 10b. The confusion matrix (CM) for Xception model is also calculated to help in understanding the results. It displays the number of true positives (TP), false positives (FP), true negatives (TN), and false negatives (FN) as shown in figure10c. It is inferred from the CM that FP and FN have increased in comparison to the ViT model, thus showing lesser correct predictions of images into normal and OSCC classes. After training and validation, we obtained the ROC (receiver operating characteristic) curve for the model, which shows performance exceeding 0.5 and an AUC (area under the curve) value of 0.89 as shown in figure 11a. The precision-recall curve in figure 11b shows an average precision of 0.85.

**Figure 10:**
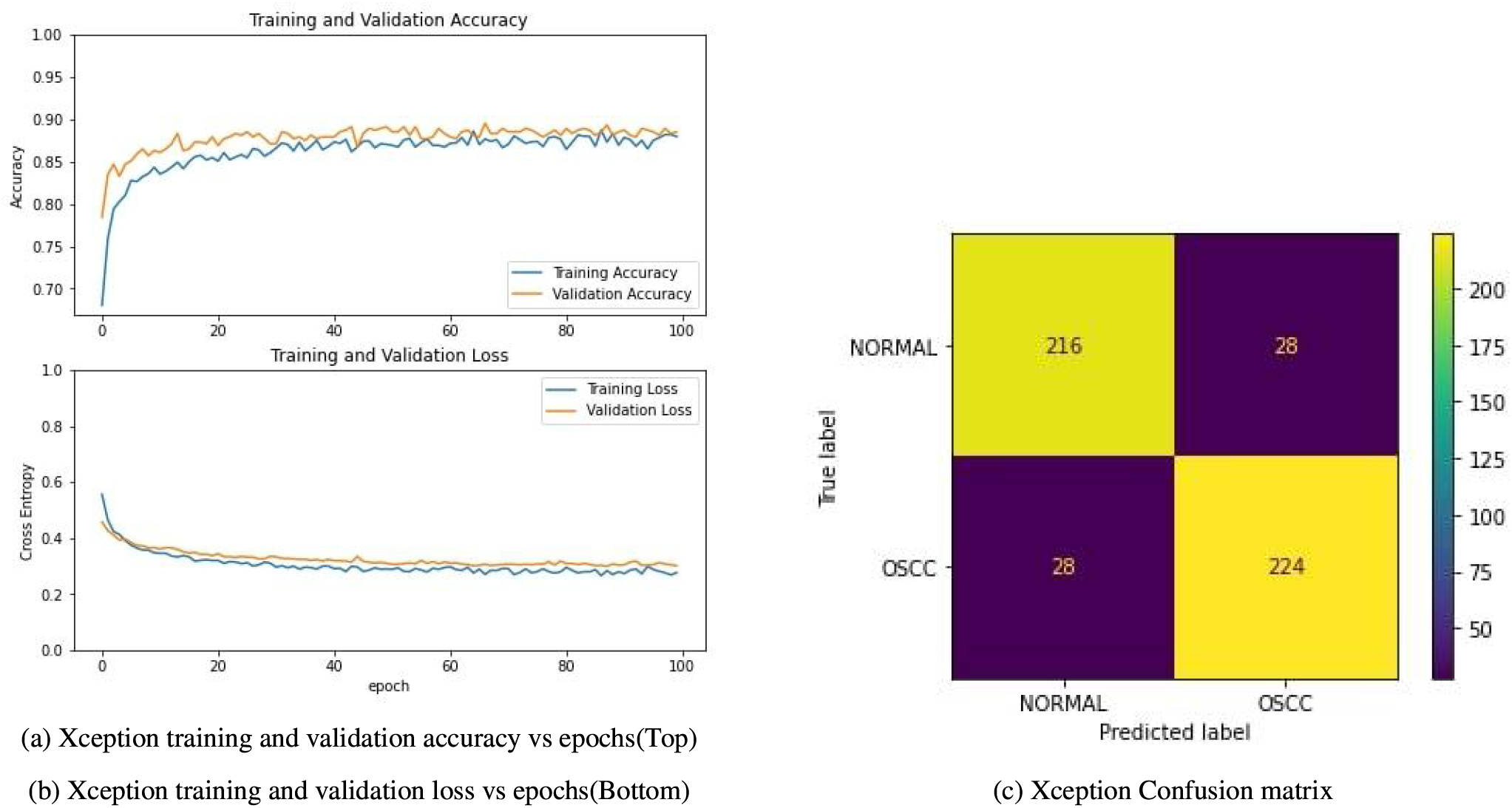
(a)Accuracy plot of Xception model training over 100 epochs(b)Loss plot of Xception model training over 100 epochs(c)Confusion matrix shows that there are 216 true negatives, 28 false positives, 28 false negatives and 224 true positives

**Figure 11:**
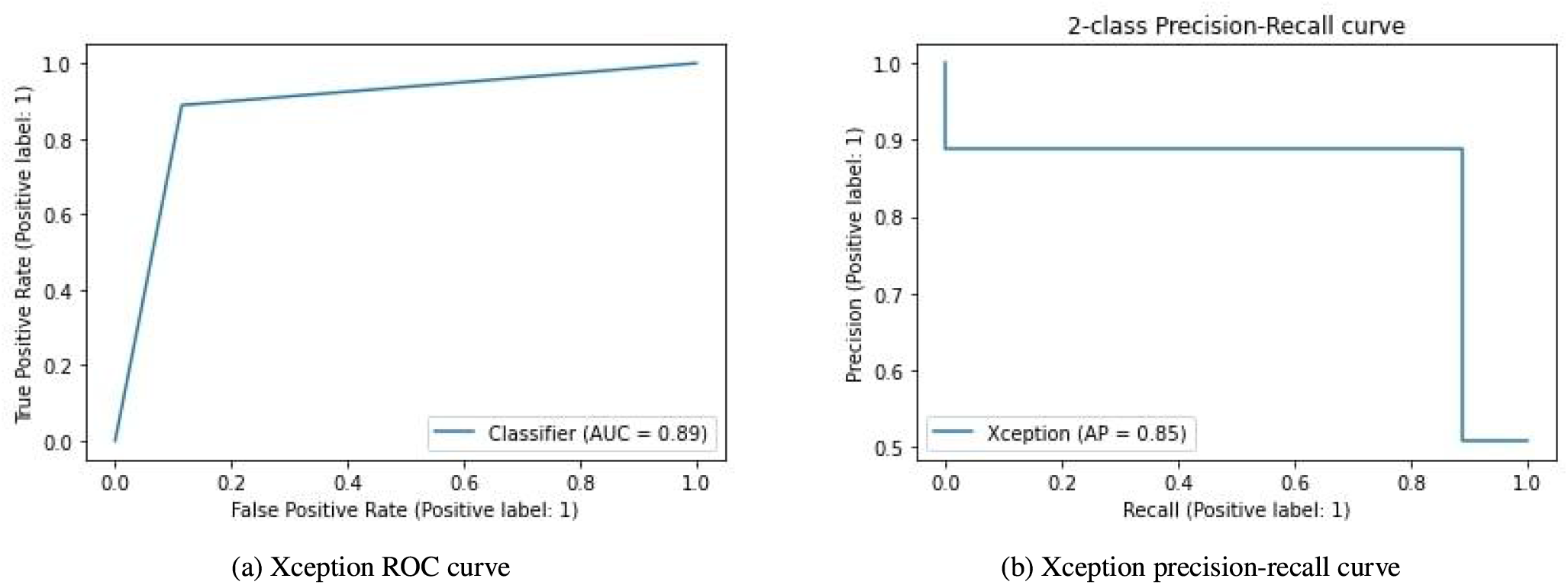
Receiver Operating Characteristic curve for the Xception model. Area under the curve is calculated as 0.89(b)The precision-recall curve is constructed by plotting the precision against the recall for the Xception model. Average precision value is 0.85 which shows that our xception model gets lesser correct predictions compared to our proposed ViT model.

#### 6.3.3. Discussion of Resnet50 model results

For the analytical results of Resnet50 model on the histopathological oral cancer dataset, we have utilized a model pre-trained on the ImageNet dataset. We have used Resnet50 as the base model and an additional softmax classifier on top of it to categorize images into normal and OSCC classes. The training and validation accuracy and loss curves are plotted over 100 epochs as displayed in figure 12a and 12b. The confusion matrix (CM) for Resnet50 model is also calculated to help in understanding the results. It displays the number of true positives (TP), false positives (FP), true negatives (TN), and false negatives (FN) as shown in figure12c. The confusion matrix shows that there are 211 true negatives, 33 false positives, 17 false negatives and 235 true positives. After training and validation, we obtained the ROC (receiver operating characteristic) curve for the model, which shows performance exceeding 0.5 and an AUC (area under the curve) value of 0.90 as shown in figure 13a. The precision-recall curve in figure 13b shows an average precision of 0.85.

**Figure 12:**
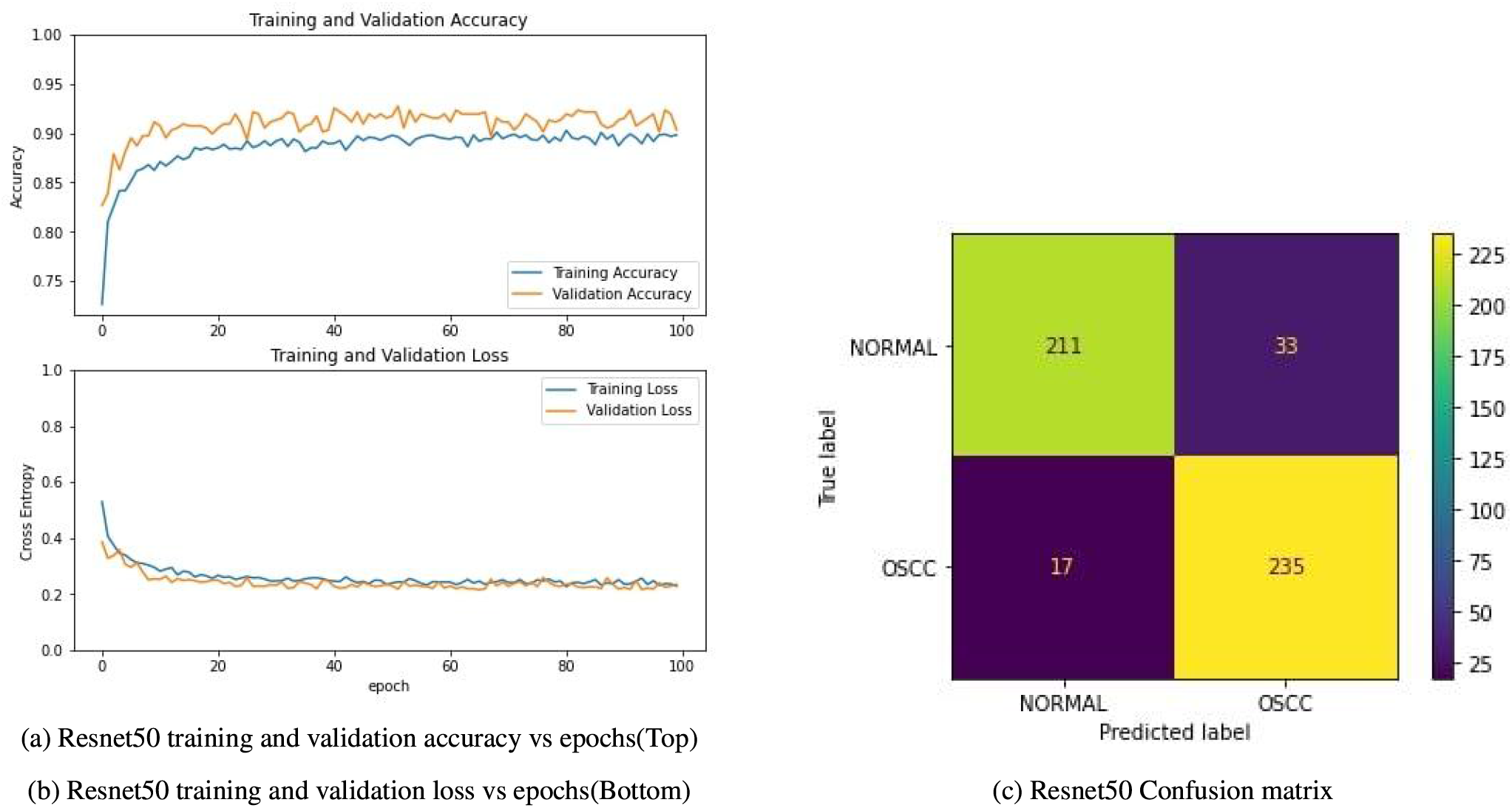
(a)Accuracy plot of Resnet50 model training over 100 epochs(b)Loss plot of Resnet50 model training over 100 epochs(c)Confusion matrix shows that there are 211 true negatives, 33 false positives, 17 false negatives and 235 true positives

**Figure 13:**
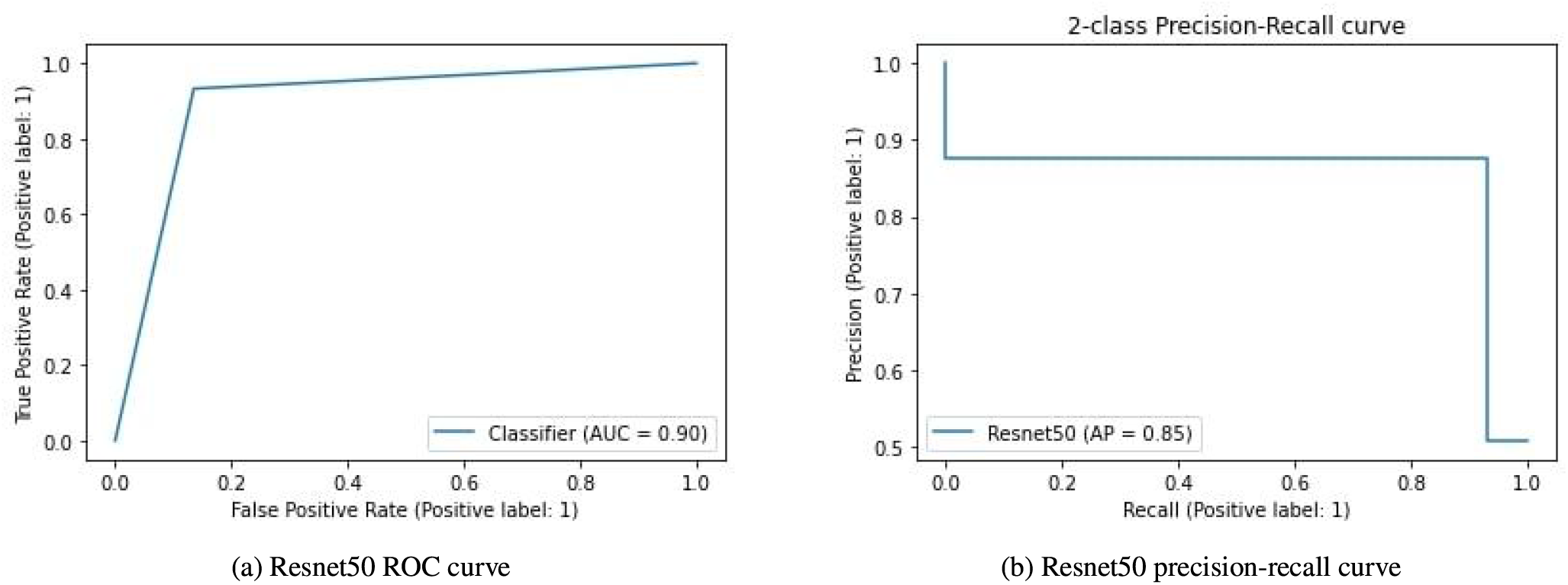
Receiver Operating Characteristic curve for the Resnet50 model. Area under the curve is calculated as 0.90 (b)The precision-recall curve is constructed by plotting the precision against the recall for the Resnet50 model. Average precision value is 0.85 which shows that our Resnet50 model gets lesser correct predictions compared to our proposed ViT model.

#### 6.3.4. Discussion of Inceptionv3 model results

For the analytical results of InceptionV3 model on the histopathological oral cancer dataset, we have utilized a model pre-trained on the ImageNet dataset. We have used InceptionV3 as the base model and an additional softmax classifier on top of it to categorize images into normal and OSCC classes. The training and validation accuracy and loss curves are plotted over 100 epochs as displayed in figure 14a and 14b. The confusion matrix (CM) for InceptionV3 model is also calculated to help in understanding the results. It displays the number of true positives (TP), false positives (FP), true negatives (TN), and false negatives (FN) as shown in figure 14c. The confusion matrix shows that there are 200 true negatives, 44 false positives, 35 false negatives, and 217 true positives. After training and validation, we obtained the ROC (receiver operating characteristic) curve for the model, which shows performance exceeding 0.5 and an AUC (area under the curve) value of 0.84 as shown in figure 15a. The precision-recall curve in figure 15b shows an average precision of 0.79.

**Figure 14:**
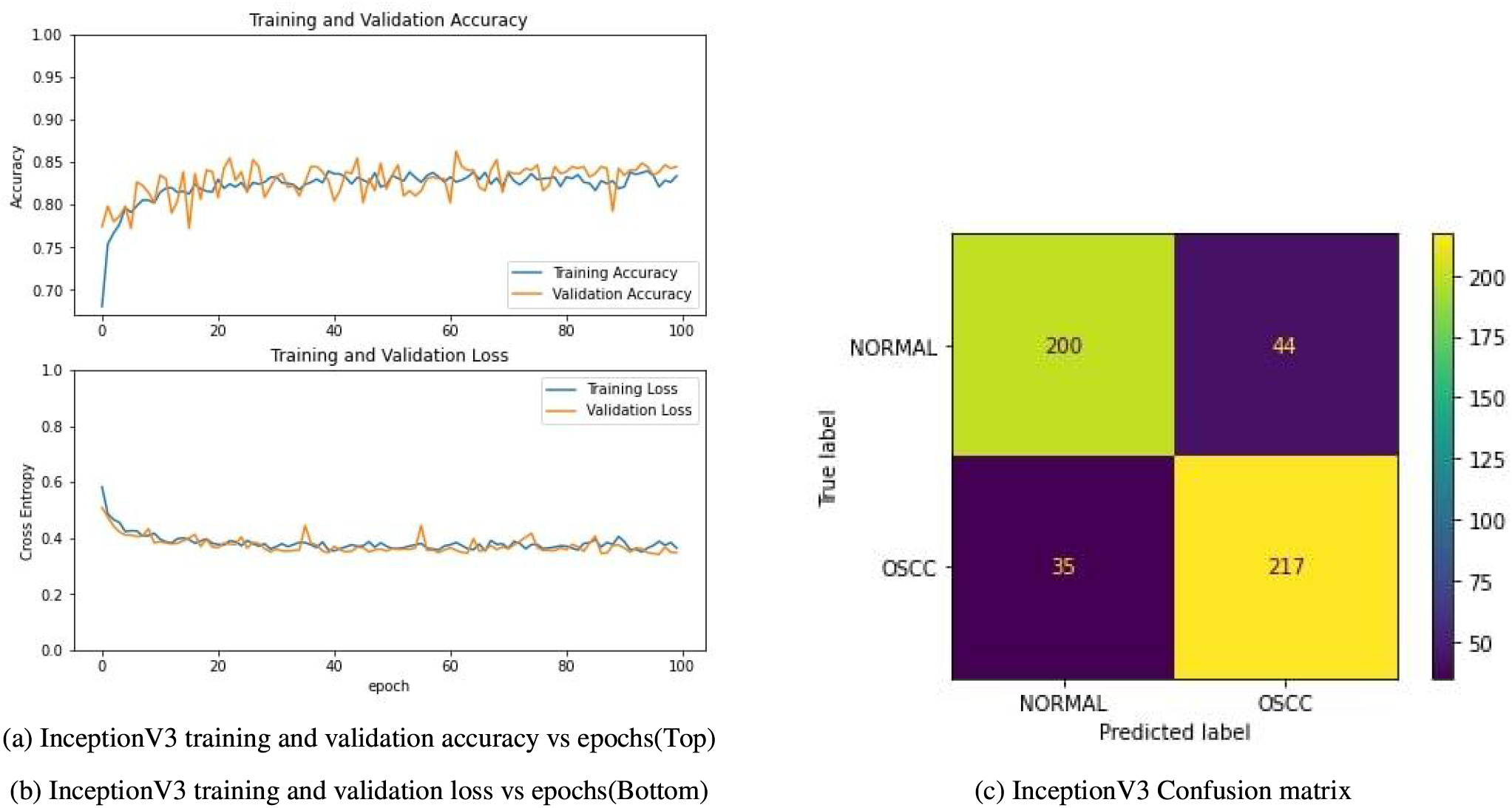
(a)Accuracy plot of InceptionV3 model training over 100 epochs(b)Loss plot of InceptionV3 model training over 100 epochs(c)Confusion matrix shows that there are 200 true negatives, 44 false positives, 35 false negatives and 217 true positives

**Figure 15:**
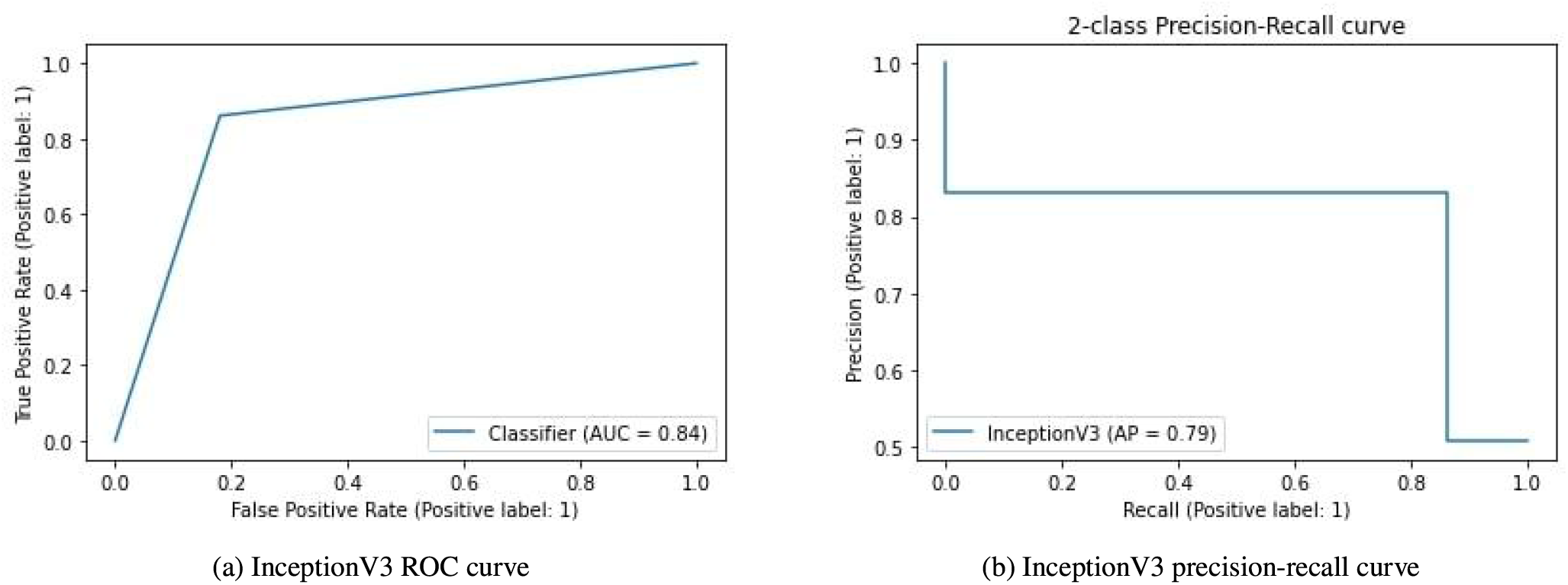
Receiver Operating Characteristic curve for the InceptionV3 model. Area under the curve is calculated as 0.84 (b)The precision-recall curve is constructed by plotting the precision against the recall for the InceptionV3 model. Average precision value is 0.79 which shows that our InceptionV3 model gets lesser correct predictions compared to our proposed ViT model.

#### 6.3.5. Discussion of InceptionResnetV2 model results

For the analytical results of InceptionResnetV2 model on the histopathological oral cancer dataset, we have utilized a model pre-trained on the ImageNet dataset. We have used InceptionResnetV2 as the base model and an additional softmax classifier on top of it to categorize images into normal and OSCC classes. The training and validation accuracy and loss curves are plotted over 100 epochs as displayed in 16a and 16b. The confusion matrix (CM) for InceptionResnetV2 model is also calculated to help in understanding the results. It displays the number of true positives (TP), false positives (FP), true negatives (TN), and false negatives (FN) as shown in figure 16c. The confusion matrix shows that there are 200 true negatives, 44 false positives, 22 false negatives and 230 true positives. After training and validation, we obtained the ROC (receiver operating characteristic) curve of the model, which shows performance exceeding 0.5 and an AUC (area under the curve) value of 0.87 as shown in figure 17a. The precision-recall curve in figure 17b shows an average precision of 0.81.

**Figure 16:**
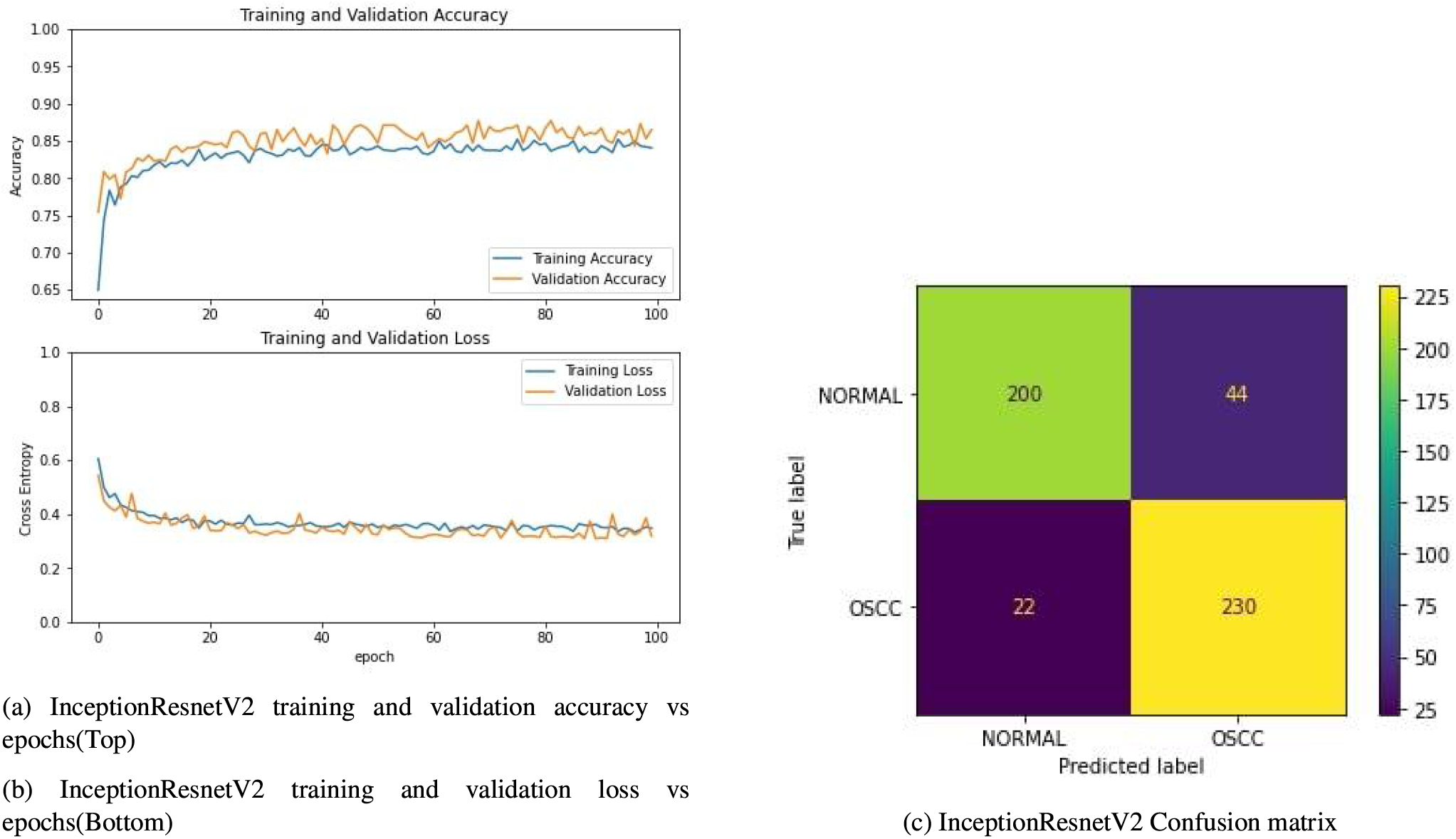
(a)Accuracy plot of InceptionResnetV2 model training over 100 epochs(b)Loss plot of InceptionResnetV2 model training over 100 epochs(c)Confusion matrix shows that there are 200 true negatives, 44 false positives, 22 false negatives and 230 true positives.

**Figure 17:**
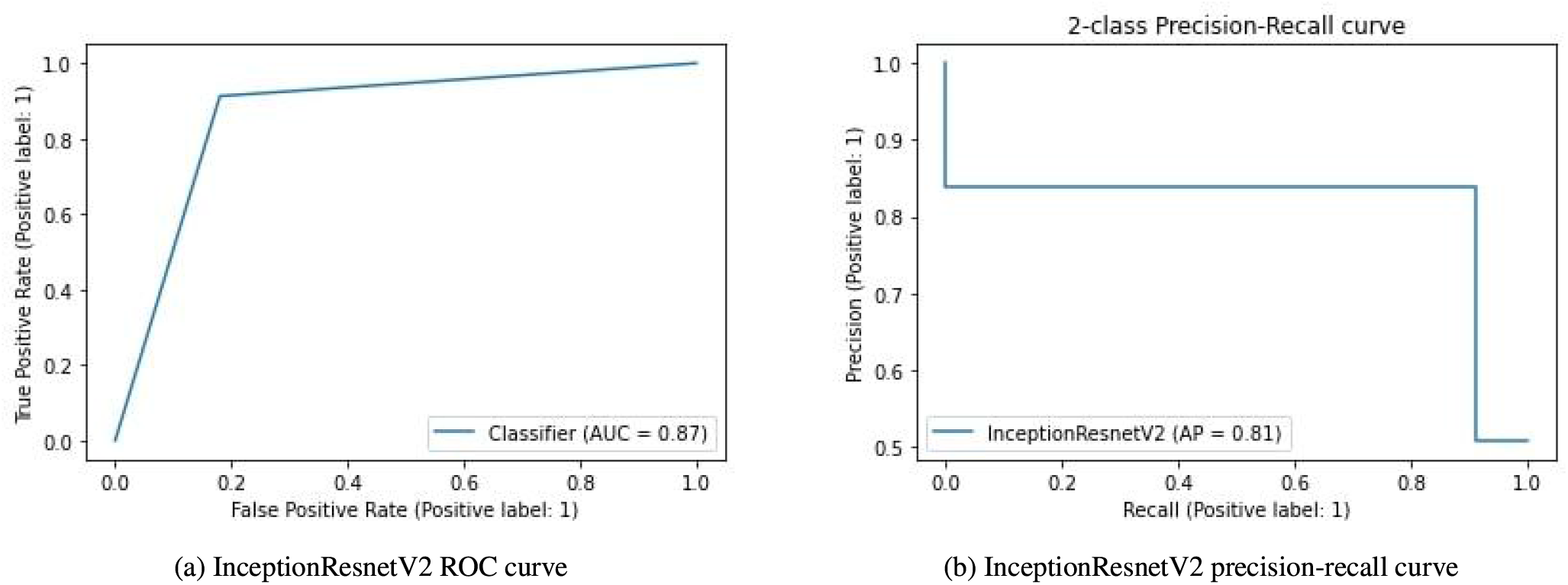
Receiver Operating Characteristic curve for the InceptionResnetV2 model. Area under the curve is calculated as 0.87 (b)The precision-recall curve is constructed by plotting the precision against the recall for the InceptionResnetV2 model. Average precision value is 0.81 which shows that our InceptionResnetV2 model gets lesser correct predictions compared to our proposed ViT model.

#### 6.3.6. Discussion of Densenet121 model results

For the analytical results of Densenet121 model on the histopathological oral cancer dataset, we have utilized a model pre-trained on the ImageNet dataset. We have used Densenet121 as the base model and an additional softmax classifier on top of it to categorize images into normal and OSCC classes. The training and validation accuracy and loss curves are plotted over 100 epochs as displayed in figure 18a and 18b. The confusion matrix (CM) for Densenet121 model is also calculated to help in understanding the results. It displays the number of true positives (TP), false positives (FP), true negatives (TN), and false negatives (FN) as shown in figure 18c. The confusion matrix shows that there are 168 true negatives, 76 false positives, 14 false negatives, and 238 true positives. After training and validation, we obtained the ROC (receiver operating characteristic) curve for the model, which shows performance exceeding 0.5 and an AUC (area under the curve) value of 0.82 as shown in figure 19a. The precision-recall curve in figure 19b shows an average precision of 0.74.

**Figure 18:**
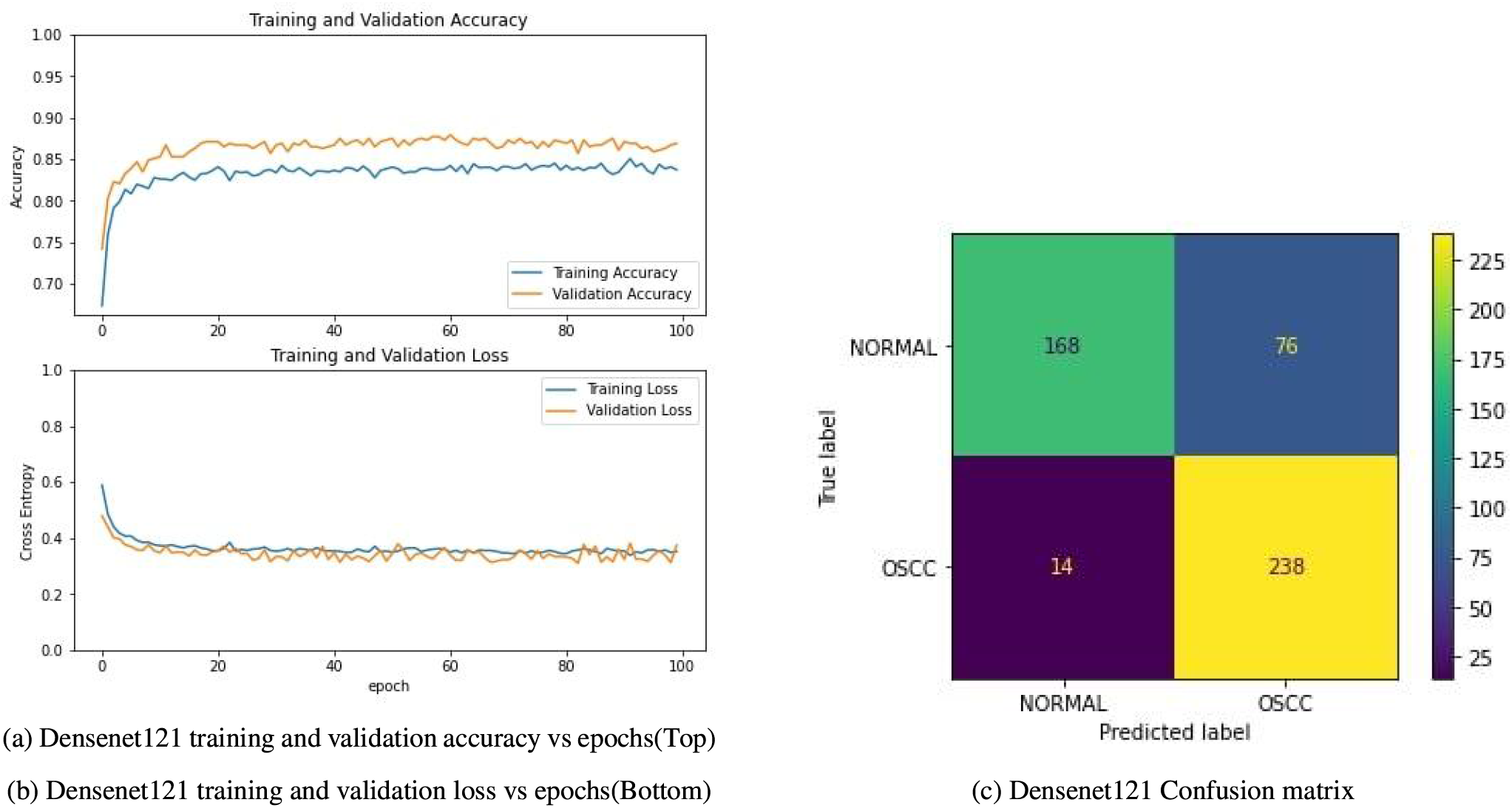
(a)Accuracy plot of Densenet121 model training over 100 epochs(b)Loss plot of Densenet121 model training over 100 epochs(c)Confusion matrix shows that there are 168 true negatives, 76 false positives, 14 false negatives and 238 true positives.

**Figure 19:**
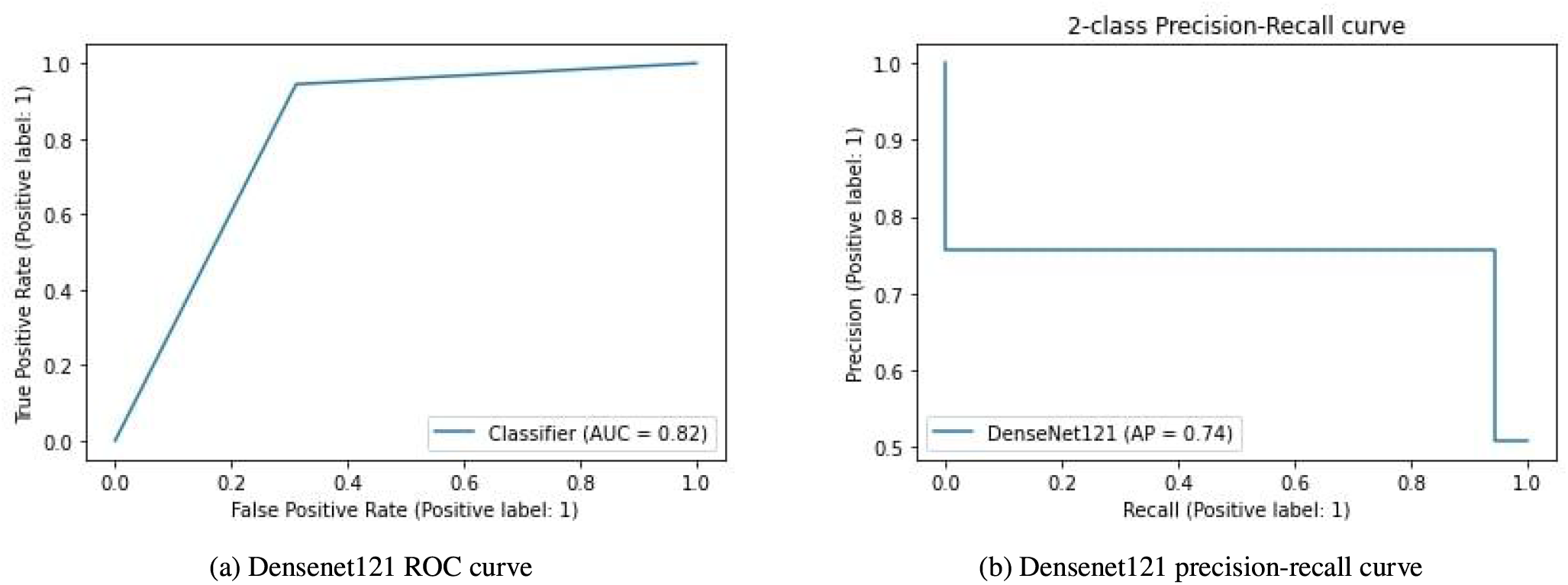
Receiver Operating Characteristic curve for the Densenet121 model. Area under the curve is calculated as 0.82 (b)The precision-recall curve is constructed by plotting the precision against the recall for the Densenet121 model. Average precision value is 0.74 which shows that our Densenet121 model gets lesser correct predictions compared to our proposed ViT model.

#### 6.3.7. Discussion of Densenet169 model results

For the analytical results of Densenet169 model on the histopathological oral cancer dataset, we have utilized a model pre-trained on the ImageNet dataset. We have used Densenet169 as the base model and an additional softmax classifier on top of it to categorize images into normal and OSCC classes. The training and validation accuracy and loss curves are plotted over 100 epochs as displayed in figure 20a and 20b. The confusion matrix (CM) for Densenet169 model is also calculated to help in understanding the results. It displays the number of true positives (TP), false positives (FP), true negatives (TN), and false negatives (FN) as shown in figure 20c. The confusion matrix shows that there are 187 true negatives, 57 false positives, 19 false negatives, and 233 true positives. After training and validation, we obtained the ROC (receiver operating characteristic) curve of the model, which shows performance exceeding 0.5 and an AUC (area under the curve) value of 0.85 as shown in figure 21a. The precision-recall curve in figure 21b shows an average precision of 0.78.

**Figure 20:**
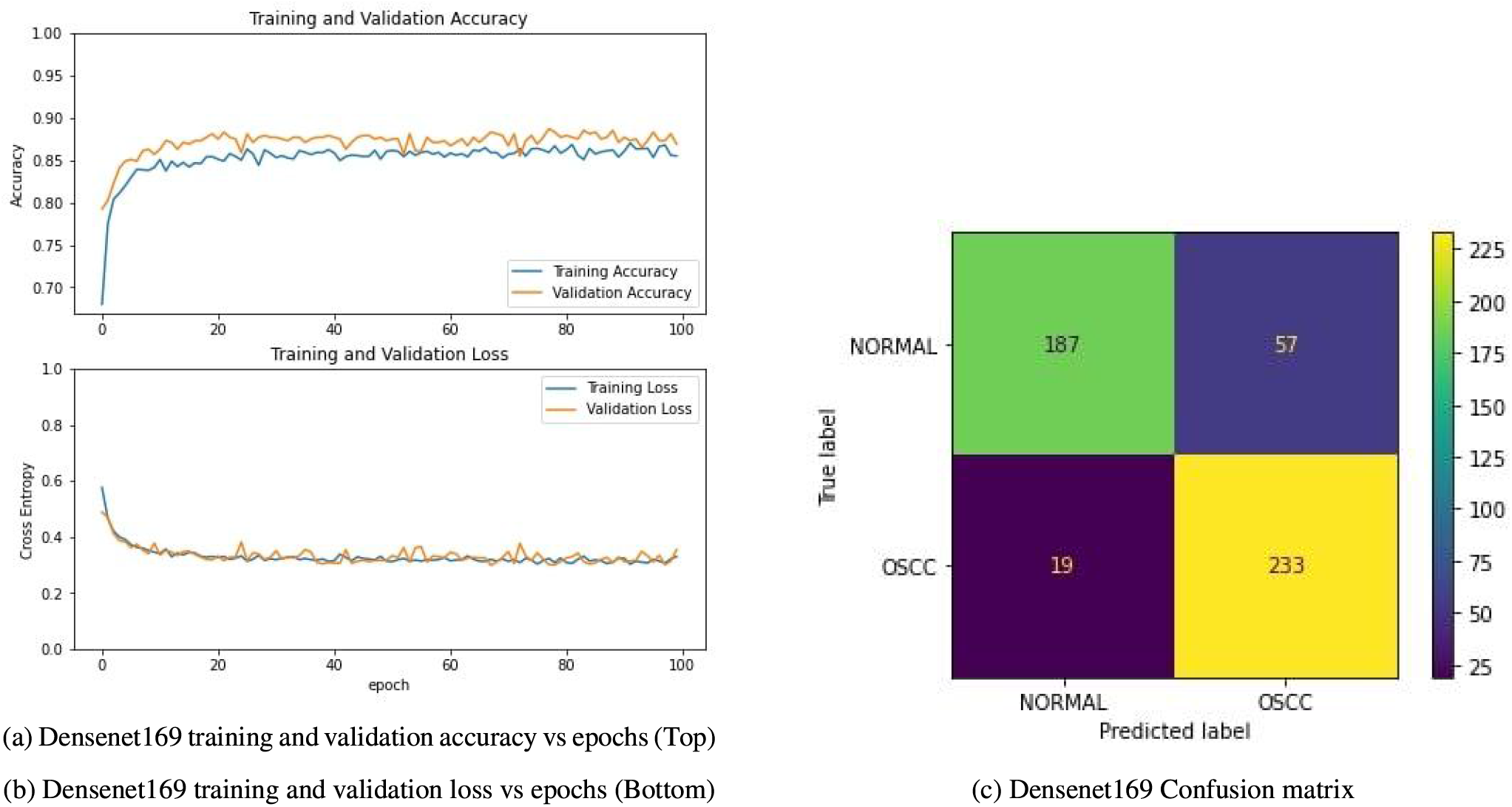
(a)Accuracy plot of Densenet169 model training over 100 epochs(b)Loss plot of Densenet169 model training over 100 epochs(c)Confusion matrix shows that there are 187 true negatives, 57 false positives, 19 false negatives and 233 true positives.

**Figure 21:**
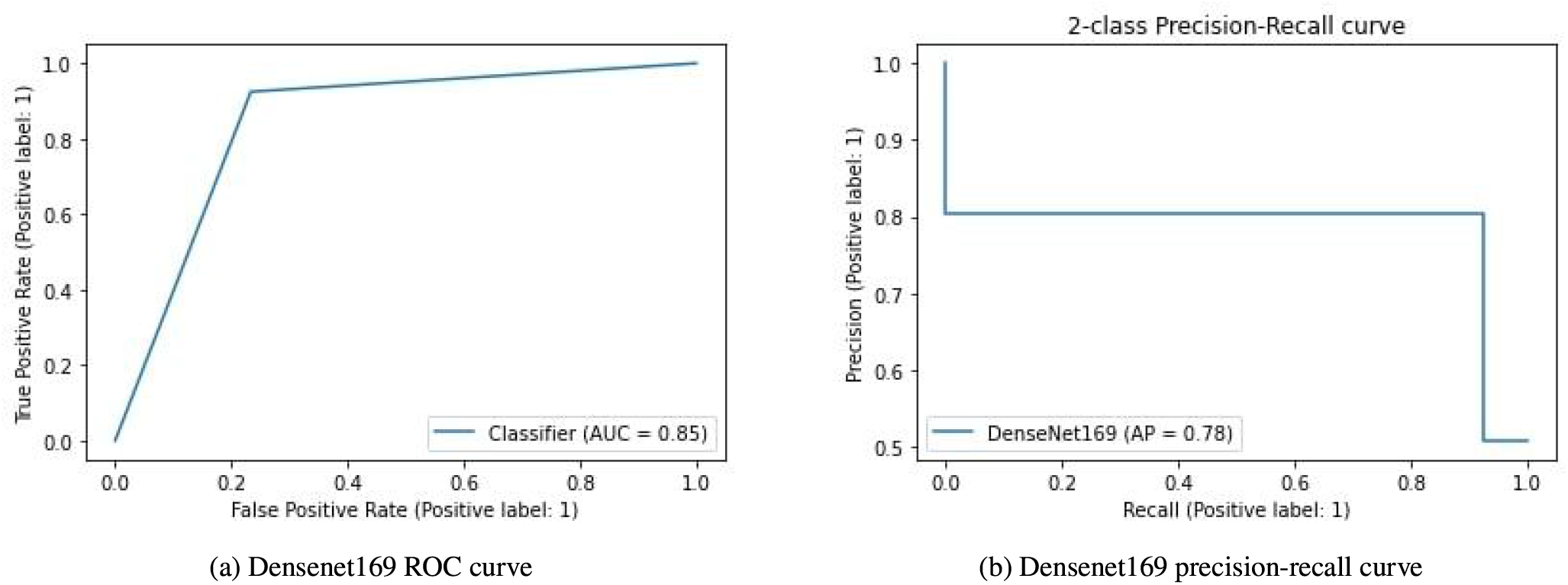
Receiver Operating Characteristic curve for the Densenet169 model. Area under the curve is calculated as 0.85 (b)The precision-recall curve is constructed by plotting the precision against the recall for the Densenet169 model. Average precision value is 0.78 which shows that our Densenet169 model gets lesser correct predictions compared to our proposed ViT model.

#### 6.3.8. Discussion of Densenet201 model results

For the analytical results of Densenet201 model on the histopathological oral cancer dataset, we have utilized a model pre-trained on the ImageNet dataset. We have used Densenet201 as the base model and an additional softmax classifier on top of it to categorize images into normal and OSCC classes. The training and validation accuracy and loss curves are plotted over 100 epochs as displayed in figure 22a and 22b. The confusion matrix (CM) for Densenet201 model is also calculated to help in understanding the results. It displays the number of true positives (TP), false positives (FP), true negatives (TN), and false negatives (FN) as shown in figure 22c. The confusion matrix shows that there are 205 true negatives, 39 false positives, 17 false negatives, and 235 true positives. After training and validation, we obtained the ROC (receiver operating characteristic) curve for the model, which shows performance exceeding 0.5 and an AUC (area under the curve) value of 0.89 as shown in figure 23a. The precision-recall curve in figure 23b shows an average precision of 0.83.

**Figure 22:**
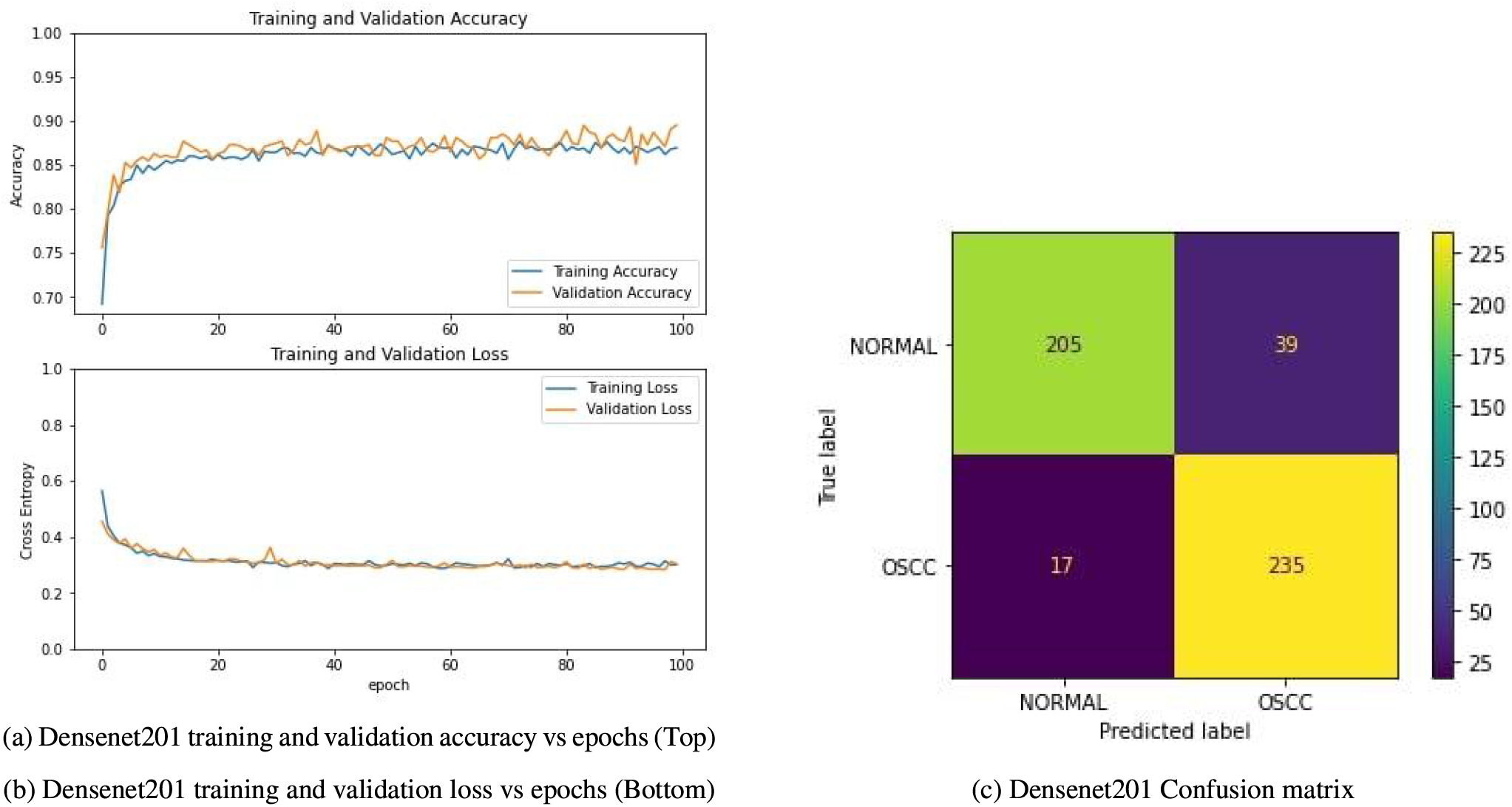
(a)Accuracy plot of Densenet201 model training over 100 epochs(b)Loss plot of Densenet201 model training over 100 epochs(c)Confusion matrix shows that there are 205 true negatives, 39 false positives, 17 false negatives and 235 true positives.

**Figure 23:**
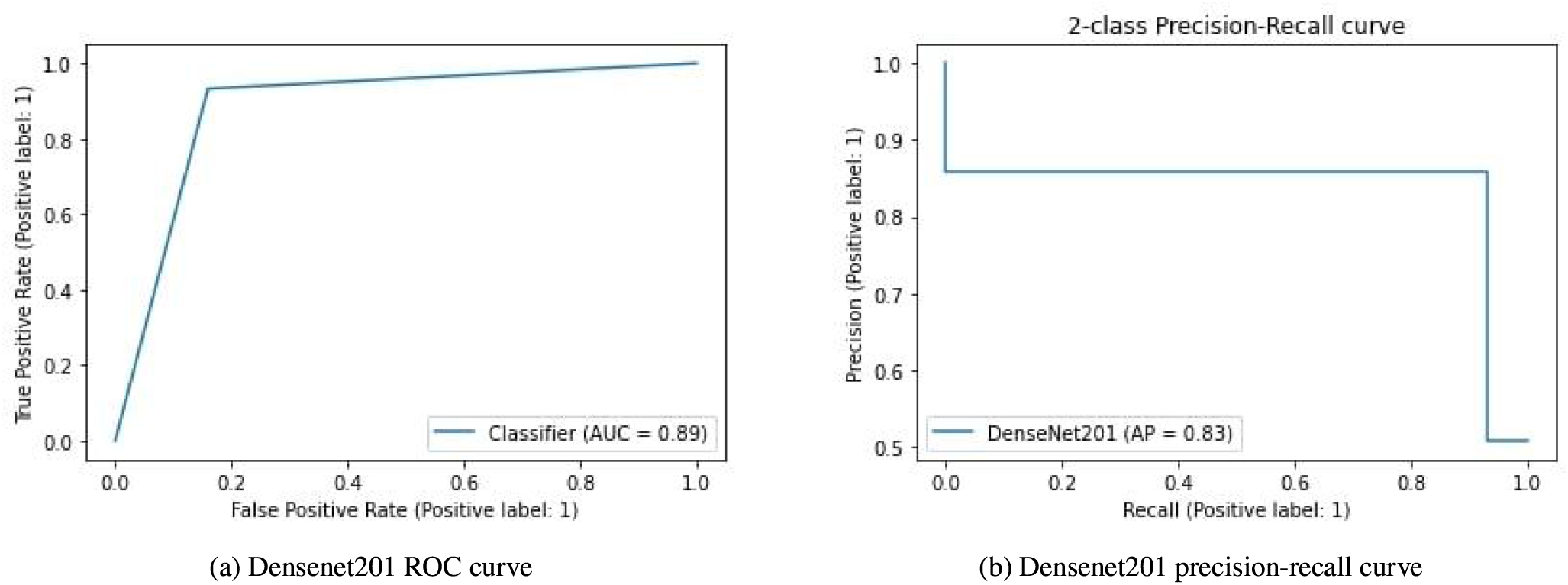
Receiver Operating Characteristic curve for the Densenet201 model. Area under the curve is calculated as 0.89 (b)The precision-recall curve is constructed by plotting the precision against the recall for the Densenet201 model. Average precision value is 0.83 which shows that our Densenet201 model gets lesser correct predictions compared to our proposed ViT model.

#### 6.3.9. Discussion of EfficientNetB7 model results

For the analytical results of EfficientNetB7 model on the histopathological oral cancer dataset, we have utilized a model pre-trained on the ImageNet dataset. We have used EfficientNetB7 as the base model and an additional softmax classifier on top of it to categorize images into normal and OSCC classes. The training and validation accuracy and loss curves are plotted over 100 epochs as displayed in figure 24a and 24b. The confusion matrix (CM) for EfficientNetB7 model is also calculated to help in understanding the results. It displays the number of true positives (TP), false positives (FP), true negatives (TN), and false negatives (FN) as shown in figure 24c. The confusion matrix shows that there are 203 true negatives, 41 false positives, 22 false negatives, and 230 true positives. After training and validation, we obtained the ROC (receiver operating characteristic) curve of the model, which shows performance exceeding 0.5 and an AUC (area under the curve) value of 0.87 as shown in figure 25a. The precision-recall curve in figure 25b shows an average precision of 0.82.

**Figure 24:**
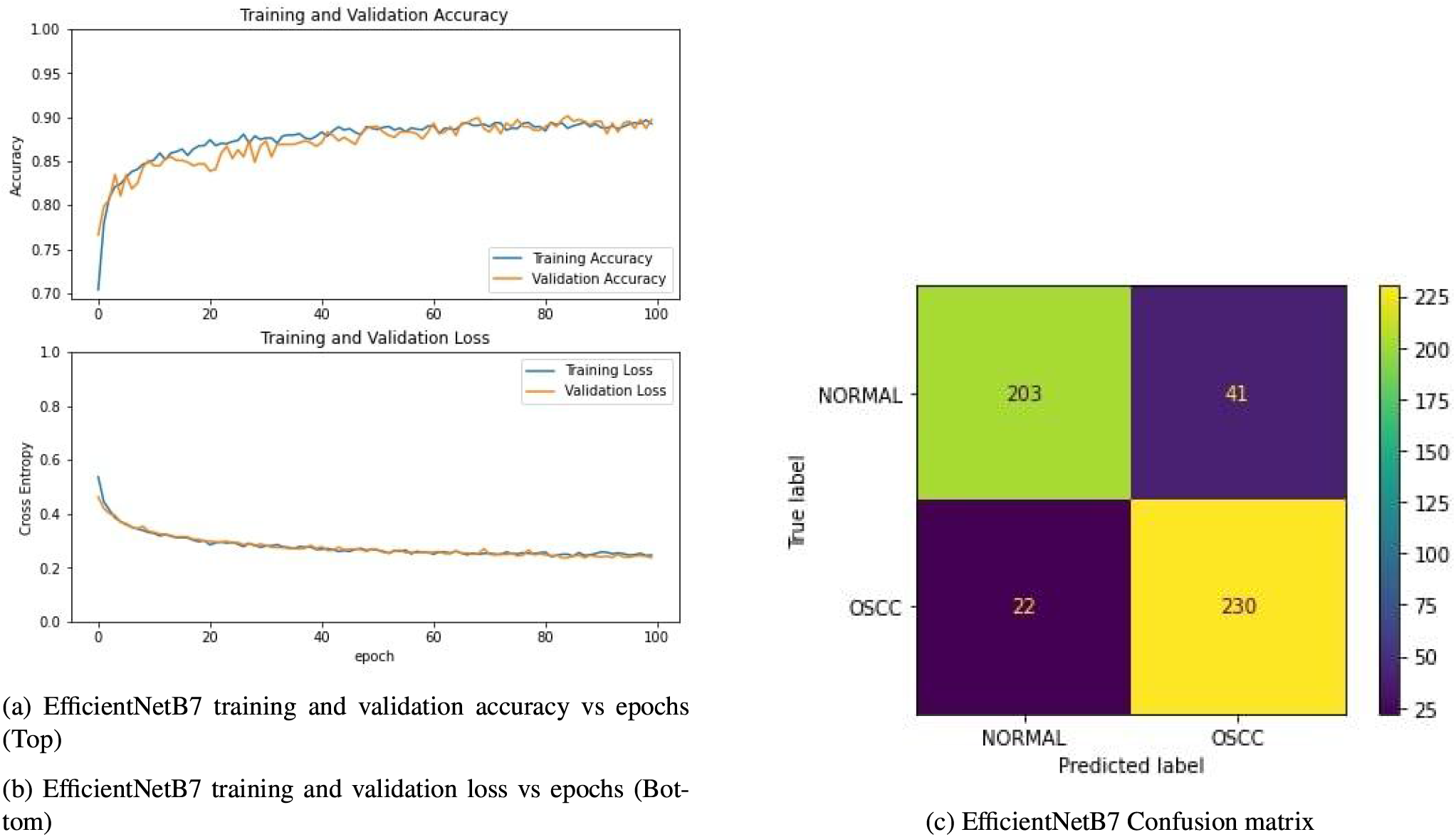
(a)Accuracy plot of EfficientNetB7 model training over 100 epochs(b)Loss plot of EfficientNetB7 model training over 100 epochs(c)Confusion matrix shows that there are 203 true negatives, 41 false positives, 22 false negatives and 230 true positives.

**Figure 25:**
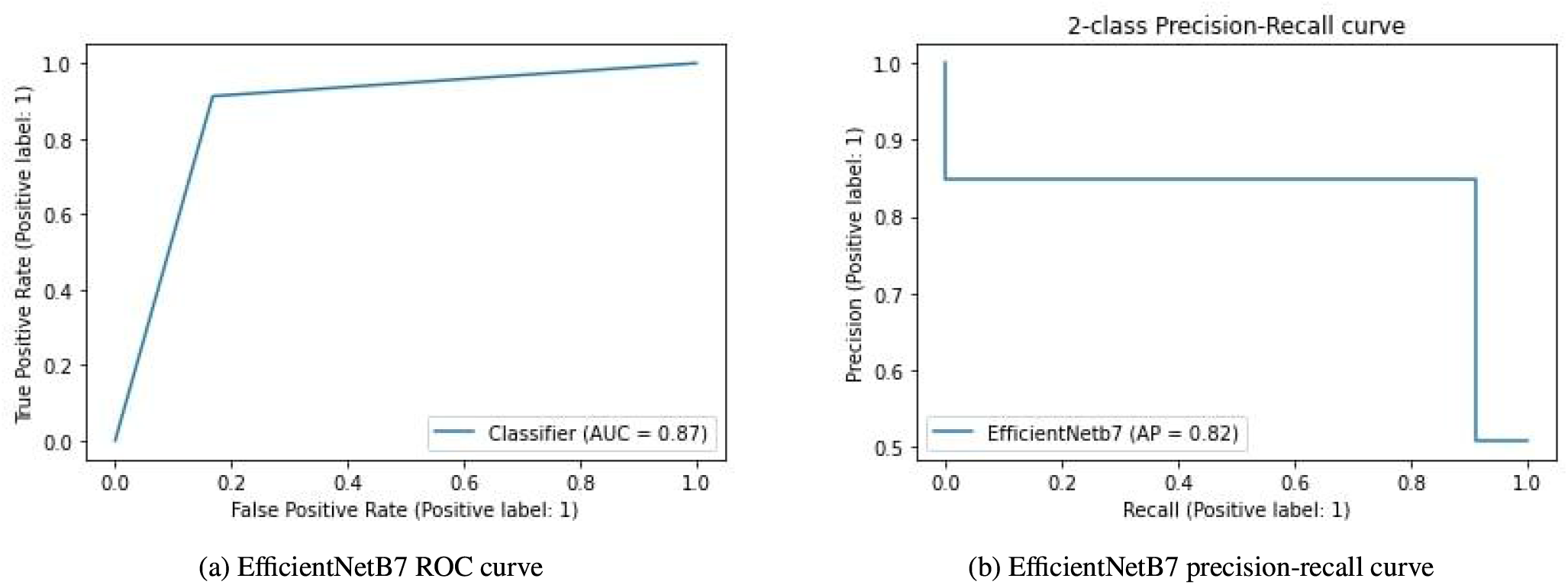
Receiver Operating Characteristic curve for the EfficientNetB7 model. Area under the curve is calculated as 0.87 (b)The precision-recall curve is constructed by plotting the precision against the recall for the EfficientNetB7 model. Average precision value is 0.82 which shows that our EfficientNetB7 model gets lesser correct predictions compared to our proposed ViT model.

## 7. Conclusion

In this paper, transfer learning of transformers for histopathological images are assessed. The classification of oral cancer images is done using a transformer-based approach. Kaggle’s Oral Cancer dataset is utilised to evaluate performance. Due to its superior performance and enormous potential in comparison to deep learning (DL) models, Vision Transformers (ViT) are now among the popular topics in the field of computer vision. Although DL are sufficiently developed to support the expansion of applications that can guarantee an effective and accurate medical assessment. However, the idea of attention in Vision Transformers has made its way for more precise results in the medical industry, where an inaccurate output could harm lives. ViT models perform more precisely than DL models because they evaluate the image’s overall context as well as its interpretability via attention module.

Our approach is based on transfer learning strategies as well as the transformer’s original architecture. The image classification model makes use of the encoder block of the transformer. A combination of new models as well as a CNN and transformer architecture may be used in future model efficacy assessments for medical imaging. In terms of overall accuracy, the proposed Vision Transformer model achieved a binary classification accuracy performance of 97.78%. Based on the used evaluation criteria, the suggested model outperforms state-of-the-art deep learning algorithms in binary classification, demonstrating its superiority and effectiveness in the early detection of oral cancer using histopathological images.

The key disadvantage of this strategy is that if the data are scarce, it may be challenging to establish its robustness and generalizability (61). Only 4946 images make up the dataset used in this work, although larger datasets are needed to train more effective deep learning models, depending on how difficult the problem is. Fundamentally speaking, a very large database is optimal for deep learning models. We used transfer learning models that were pretrained on ImageNet and then refined using the histopathological images in our work because ImageNet has 14 million images. Future directions for this study include expanding our dataset and incorporating images for multi-class classification problems. We will evaluate ViT model applications using histopathology images of different diseases, including breast, cervical, and lung cancer.

## Data Availability

https://www.kaggle.com/ashenafifasilkebede/dataset?select=val

## CRediT authorship contribution statement

**Bhaswati Singha Deo:** Developed the model from the concept, Developed the code, Generated the results, Wrote the manuscript. **Mayukha Pal:** Conceived the idea and conceptualized it, Developed the methodology from the concept, Reviewed the manuscript and mentored the work. **Prasanta K. Panigrahi:** Reviewed the manuscript and mentored the work. **Asima Pradhan:** Developed the analysis methodology, Reviewed the manuscript and mentored the work.

## Declaration of Competing Interest

The authors declare that they have no known competing financial interests or personal relationships that could have appeared to influence the work reported in this paper.

## Acknowledgement

This research received no specific support from governmental, private, or non-profit funding agencies. The content and writing of the paper are solely the responsibility of the authors. Bhaswati Singha Deo is thankful to IIT Kanpur for the Institute fellowship. The author MPal wishes to thank ABB Ability Innovation Centre, India for their support in this work.

## Notes

### Competing Interest Statement

The authors have declared no competing interest.

### Funding Statement

This study did not receive any funding

